# Changes in household food security, access to health services, and income in northern Lao PDR during the COVID-19 pandemic

**DOI:** 10.1101/2021.07.27.21261221

**Authors:** Jennifer R. Head, Phetsavanh Chanthavilay, Helen Catton, Ammaline Vongsitthi, Kelley Khamphouxay, Niphone Simphaly

## Abstract

**Background:** The COVID-19 pandemic is expected to exacerbate food insecurity in low- and middle-income countries, through loss of income and disrupted food supply chains. Lao PDR has among the highest rates of malnutrition in Southeast Asia. We assessed the relative difficulty in meeting food needs during the COVID-19 pandemic in rural districts of Luang Prabang Province, Lao PDR compared to before; determined associations between pandemic-associated difficulties in food access and household, maternal and child food security; and identified resiliency-promoting strategies.

**Methods:** In November 2020, households (N = 1,122) with children under five years were interviewed. Respondents reported the relative ease of access of food and health care as well as changes in income and expenditures compared to before March 2020. We used generalized linear models with cluster robust standard errors to assess univariate and multivariate associations.

**Results:** Nearly four-fifths (78.5%) found it harder to meet household food needs during the pandemic. The most common reasons were increased food prices (51.2%), loss of income (45.3%), and decreased food availability (36.6%). Adjusting for demographics, households with increased difficulty meeting food needs had lower food consumption scores and child dietary diversity. Over 85% of households lost income during the pandemic. Decreased expenditures was associated with reliance on more extreme coping strategies to meet food needs. The households who experienced no change in meeting food needs produced a greater percentage of their food from homegrown methods (4.22% more, 95% CI: 1.28, 7.15), than households who found it more difficult. We estimated that decreases in child bodyweight by 0.5 – 1% would increase wasting in this population by 1.7 – 2.1 percentage points.

**Conclusions:** Pandemic-associated shocks may have large effects on malnutrition prevalence. Action is needed to mitigate consequences of the pandemic on nutrition. Local food production and safety net programs that offset income losses may help.

**Summary Box:** *What is already known?:* The COVID-19 pandemic has disrupted food supply chains and livelihoods, causing concerns that a global nutrition crisis is imminent and prompting leaders from United Nations agencies to issue an immediate call to action to direct funds towards prevention of child malnutrition. While documented COVID-19 transmission in Lao PDR was lower than that of surrounding counties, malnutrition rates are high, particularly in the northern province of Luang Prabang, which is heavily reliant on tourism for livelihoods.

*What are the new findings?:* Nearly four-fifths of those interviewed in Luang Prabang Province, Lao PDR reported that it was harder to meet their household’s food needs, compared to before the pandemic, with 51% attributing the reason to increased food prices. Over 85% of households reported losing income. Lower expenditures and increased difficulty obtaining food were both associated with lower household food consumption scores and higher household coping strategies, in adjusted analyses. Households who obtained a greater proportion of their foods through home production appeared more resilient than households who obtained a greater proportion of their foods through purchasing.

*What do the new findings imply?:* The pandemic may deeply exacerbate food insecurity in Lao PDR, potentially leading to increases in child wasting. Increased local food production and establishment of safety net programs that offset income losses may be two strategies that address this problem among this population.

## Introduction

Disruptions to food, economics, and health systems during the COVID-19 pandemic are expected to increase the risk of malnutrition among low- and middle-income countries (LMICs) [1–4]. The food supply chain has faced challenges across multiple stages, including loss of labor for agricultural production and postharvest handling due to movement restrictions or illnesses; closure of processing and distributing facilities; disruptions in distribution networks under restricted trade policies; and changes in consumer demand and market access [5]. Such challenges have resulted in increases in food prices, with the Food and Agricultural Organization (FAO) reporting that wheat and rice prices increased by 8% and 25%, respectively, between March 2019 and April 2020 [6]. Economic disruptions, such as business closures and declines in tourism, are simultaneously expected to reduce country-specific gross national incomes (GNI) by around 8% in most LMICs [7]. Losses in income are expected to push an additional 1.4 million people into extreme poverty, classified as earning less than $1.90 per day [7]. Overall, the World Food Programme projects that the number of people in LMICs who are food insecure will double, from 135 million in 2019 to 265 million by the end of 2020 [8]. Compounding this effect, health services designed to catch and treat acute malnutrition may be disrupted in many LMICs. For instance, UNICEF estimates a reduction of 30% in the coverage of essential nutrition services in LMICs due to difficulties in mobility of both users and providers, interruption of non-COVID-19 services in communities, higher burdens on the health care workers, and limited personal protective equipment [9].

Increased food insecurity coupled with a decline in access to essential nutritional services is expected to lead to increases in the prevalence of childhood wasting, an acute form of malnutrition associated with elevated risk of mortality [10, 11]. One study estimates that there could be a 14.3% increase in the prevalence of moderate or severe wasting among children younger than five years in the 118 LMICs due to COVID-19-related income losses [2]. By another projection, an increase in wasting of this order of magnitude (10-50%), coupled with a decline in maternal and child health services by 9.8-15.9%, would be associated with an increase of 9.8-44.7% in under-five deaths per month [12]. To prevent a global malnutrition crisis, leaders from four United Nations agencies (UNHCR, UNICEF, FAO, WHO) have issued an immediate call to action, recommending $2.4 billion be directed to avoiding child malnutrition through wasting treatment and prevention, vitamin A supplementation, and breastfeeding support [13]. Alongside these efforts, leaders have called for research that estimates the scale and reach of nutrition challenges, including country-specific estimates of the effect of the pandemic on incomes, and the ability to meet food needs and access health services.

Lao People’s Democratic Republic (PDR) has one of the highest rates of malnutrition in southeast Asia, with a national prevalence of stunting of 33%, underweight of 21% and wasting of 9% [14]. Lao PDR experienced its first case of COVID-19 infection in March 2020 [15]. Shortly afterwards, the government imposed a strict lockdown for six weeks, stopping human movement between districts, provinces, and across the border. A total of six cases were identified between March and April 2020. Beginning in May 2020, restrictions on within-country movement eased along with adherence to protective measures (e.g., mask wearing and social distancing), but borders remain closed to everyone except those who entered the country via special mission flights, who must undergo strict quarantine and testing in government authorised facility [16]. Between March 2020 and February 2021, only 45 cases had been reported in Lao PDR, mainly among individuals returning to the country [17]. In April 2021, a second outbreak of COVID-19 occurred that spread quickly during New Year celebrations. A second lockdown was imposed on April 25^th^ with provincial and district travel restricted, surveillance on closed country borders increased, and testing and contact tracing efforts increased. Between April 1, 2021 and June 1, 2021, over 1,800 cases were confirmed, the majority in the capital city, Vientiane, with the first confirmed death from COVID-19 occurring in May of 2021 [17].

While Lao PDR has reported fewer cases of COVID-19 than its neighbouring countries, it may experience substantial economic and food security effects of the pandemic. The FAO reports that food prices in Lao PDR have increased by 7.1% between February 14, 2020 to January 30, 2021 [18]. At the same time, the Ministry of Labour and Social Welfare reported a surge in unemployment from 2% before the pandemic to 25% as of May 2020 [19]. Moreover, in a national assessment, UNICEF found that between August 2019 and August 2020, there was a 10-24% decline in the coverage of maternal health services, newborn services, routine vaccinations, screening for child wasting, and treatment of child wasting [9]. The economic effects of the pandemic are expected to be felt most strongly in Luang Prabang province, a popular tourist destination. In 2019, Luang Prabang received about 638,000 international visitors and 222,000 domestic tourists. In May 2020, 78% of Luang Prabang’s tourism enterprises were closed, and those that remained open did so largely at partial capacity [20]. This is particularly concerning, as the Luang Prabang province bears a disproportionate burden of children who are stunted (41.3%) or underweight (25%) [14].

In rural provinces of Luang Prabang where documented COVID-19 transmission was low, we aimed to 1) assess the relative difficulty in meeting food needs and accessing health care during the COVID-19 pandemic compared to before the pandemic; 2) compare self-reported difficulty in meeting food needs to indicators of food security among women, children and the household; 3) identify strategies associated with increased resiliency to food insecurity.

## Methods

### Survey region and population

We obtained data on a cross sectional, household survey from the Lao Provincial Health Department. Data were collected as part of the Lao Health Department’s endline evaluation of the Primary Health Care Program to monitor and evaluate public health activities over a three-year period, starting in 2017. Data were collected from three districts - Nan, NamBak, and Pak Ou - in Luang Prabang Province. These districts have a high prevalence of ethnic minorities, particularly Hmong and Khmu ethnicities. Livelihoods are largely agriculturally based.

### Sampling plan

The target overall sample size was 1,200 households. The sample size was chosen to detect with 95% confidence and 80% power a change from 77.7% to 83% in the proportion of women delivering with a skilled birth attendant since the baseline survey in 2017, accounting for a design effect of 1.5 and a non-response rate of 5%. A household was considered eligible for selection if members have lived in the village for at least two years, if it contained a child under the age of five, and if an adult respondent provided verbal, informed consent to participate.

Household selection followed a multistage clustered sampling design that stratified by the three districts. In the first stage, 25 villages were selected using probability proportional to size sampling. In the second stage, 30 households per village were selected using simple random sampling from a list of eligible households prepared by the village head in collaboration with the village health volunteer. The health and diet of one child under the age of five per household was assessed, and anthropometric measurements taken. If there were more than one child under five years in the house, a third stage of sampling was used, in which one child was selected using simple random sampling.

### Household questionnaire

Household questionnaires were administered verbally by trained data collectors. Information of household demographics, household food security, maternal and child diet, child anthropometrics, and self-reported changes in food access, income, expenditures and access to health services during the pandemic were collected. The survey was translated into Lao language, and back translated to ensure correct translation. One enumerator per team was also fluent in the local languages of Khmu and Hmong, in case the respondent did not speak Lao. A copy of the reduced survey tool is included in the Supplemental Info.

The endline survey used the same questionnaire as the baseline survey, which was adapted from global standard reproductive, maternal, newborn and child health and nutrition surveys, and added questions related to food security and access to health services during the pandemic. These additional questions were adapted from a standardized questionnaire developed by Save the Children, International to assess the impact of COVID-19 globally [21]. Respondents were asked if, compared to before the pandemic, it was much harder, somewhat harder, easier, or the same to meet their family’s food needs. If harder, families were asked to list the reasons why. Similarly, respondents were asked if, compared to before the pandemic, it was much harder, somewhat harder, easier, or the same to access health care. Finally, families were asked if they lost income or reduced their expenditures during the pandemic, and if so, asked to estimate by what percent.

### Calculation of household food security and maternal and child dietary diversity

Household food security was assessed through two standard indicators: the food consumption score and coping strategy index. The food consumption score (FCS) is a frequency weighted household dietary diversity score calculated by multiplying the frequency of consumption of different food groups consumed by a household during the 7 days before the survey by a weighting factor, and summing [22]. The food groups, and their respective weights include: main staples (2), pulses (3), vegetables (1), fruit (1), meat and fish (4), dairy (4), sugar (0.5), and oils/butter (0.5). Higher scores indicate better food security.

The Coping Strategies Index (CSI) was also used to compare household food security. CSI is calculated by multiplying the weekly frequency of five behaviors by the weight of the behavior and summing for all behaviors [23]. The five standard coping strategies and their severity weightings are: Eating less-preferred foods (1.0); Borrowing food/money from friends and relatives (2.0); Limiting portions at mealtime (1.0); Limiting adult intake (3.0), and reducing the number of meals per day (1.0). Lower scores indicate better food security. The CSI has good agreement with other indicators of household food insecurity, including the household food insecurity and access scale [24].

In addition, we calculated an individual dietary diversity score (DDS) for women and children aged 6-59 months [25]. DDS for children aged 24-59 months is calculated by summing the total number of food groups consumed in the previous 24 hours, where the food groups are defined as: grains, roots and white tubers; legumes and nuts; dairy products; meat; eggs; vitamin A-containing fruits and vegetables (i.e., dark-green, leafy vegetables, fruits that are orange on the inside); other fruits and vegetables. The child must consume at least four of the seven food groups to meet their minimum acceptable dietary diversity [25]. For children aged 6-23 months, breastmilk is added as an eighth food group and the child must consume five out of eight food groups to meet minimum acceptable dietary diversity.

DDS for women is tallied by adding up the number of food groups consumed out of the following ten groups: grains, roots, and white tubers; legumes; nuts and seeds; dairy products; meat; eggs; dark, leafy greens and vegetables; other vitamin-A-rich fruits and vegetables; other vegetables; other fruits. The woman must consume at least five of the ten food groups to meet her minimum dietary diversity [25]. Women who reported having an abnormal diet (i.e., ate much more or much less than normal) in the past 24 hours were excluded from analysis.

### Anthropometric analysis

Weight and height of children were recorded to the nearest 0.01 kg and 0.1 cm, respectively. Weight-for-age (WAZ), height-for-age (HAZ), and weight-for-height (WHZ) Z-scores were determined using 2006 WHO Growth Standards [26]. A child was considered stunted, wasted, or underweight if they had a WAZ, WHZ, or WAZ score below −2SD, respectively. The degree to which even small changes to body weight will translate into changes in the proportion of children classified as underweight or wasted varies between populations, as it depends on the density of Z-scores clustered around the dichotomous classification threshold of −2SD [27]. As undernutrition prevalence is a key indicator used to monitor progress and allocate nutrition and other health services, we considered the theoretical implications of increased food insecurity on undernutrition prevalence in our population. We examined the change in childhood undernutrition in our study population to a simulated reduction in bodyweight. Following prior study, we presumed potential COVID-19 associated shocks to range between a 0.5% and 1% reduction in bodyweight [27]. We simulated a reduction of 0.5% and 1% by multiplying child weight by 0.995 and 0.99, respectively, and recalculated the WAZ and WHZ scores under this simulated weight.

### Statistical analysis

Data were analyzed in R version 3.5 [28]. Survey weights were calculated using the inverse probability of selection for a child (for child outcome) or a household (for household or maternal outcomes). The survey package in R was used to calculate means and percentages accounting for survey weights, and standard errors used to calculate 95% confidence intervals were determined accounting for clustering [29]. Univariate and multivariate associations were assessed using generalized linear models, accounting for survey weights, and using cluster robust standard errors to adjust for clustering at the village level. A directed-acyclic-graph (DAG) was used to identify variables that may confound the relationship between pandemic-associated changes and household food security, where a confounder is defined as a variable associated with the exposure, causally associated with the outcome, and not on the causal pathway between exposure and outcome. Multivariate models examining the relationship between pandemic-associated changes and household food security included fixed effects for potential confounding factors of household ethnicity, household size, education level of mother and the head of household, and district. Adjusted models for maternal outcomes additionally included mother’s age, and models for children outcomes additionally included child’s age and sex.

### Ethics

Data were collected by the Lao Provincial Health Department as part of routine, non-research public health activities. We obtained data from the Lao Provincial Health Department. Ethical clearance for secondary data analysis was obtained from the Research Ethics Committee in the University of Health Sciences within the Lao Ministry of Health and Committee for the Protection of Human Subjects within University of California, Berkeley (protocol ID: 2021-05-14365).

### Public Involvement

Community members were involved in the conduct of this research. During the survey, community volunteers assisted in locating other community members for participation in the survey. Community members were informed of the results of this study during one of their monthly village health days. The results were conveyed verbally and with posters by the village health volunteers.

## Results

Interviews were completed for 1,122 households, corresponding to a 93.5% response rate. Reasons for non-response included empty house (53.8%), parent not at home (38.5%) and inaccessible house (5.1%). The most common ethnicities of those interviewed were Khmu (463, 41.3%), Lao Lom (340, 30.3%), and Hmong (281, 25.0%). Undernutrition among children under five years in the study region was high, with the survey-weighted prevalence of wasting at 4.5% (95% CI: 3.5, 5.8), underweight at 18.2% (95% CI: 15.9, 20.7%), and stunting at 32.9% (95% CI: 29.6, 36.4%).

### Food security

Nearly four-fifths (78.5%) of the study population reported that it was harder to meet their family’s food needs during the pandemic, as compared to before (Table 1). A weighted 60.9% (95% CI: 57.6, 64.1%) of individuals reported that it was somewhat harder to meet food needs, while 17.6% (95% CI: 15.4, 20.0%) reported that it was much harder. Among the 874 individuals who found it harder to meet food needs, the most common reason reported was that foods were more expensive (51.2%), followed by household losing income (45.3%), food not available at markets (36.6%), and markets being closed (36.5%). The median monthly expenditure among households was US$133. Households spent, on average, 40% of their income on food, which was increased from 30% in 2017.

**Table 1.** Self-reported effects of the pandemic on household access to food, health care, and income.

|  | Weighted percentage<br>(95% Confidence interval) | N |
| --- | --- | --- |
| **Relative ability to meet family's food needs now compared to before the pandemic (N = 1120)** |  |  |
| Easier | 0.83 (0.38, 1.82) | 8 |
| No change | 20.7 (18.3, 23.3) | 238 |
| Somewhat harder | 60.9 (57.6, 64.1) | 698 |
| Much harder | 17.6 (15.4, 20.0) | 176 |
| **Reasons it is harder to meet food needs during the pandemic (N = 874)** |  |  |
| Items more expensive | 51.2 (46.4, 56.0) | 415 |
| Household lost income | 45.3 (40.9, 49.9) | 465 |
| Less food is available | 36.6 (33.1, 40.2) | 561 |
| Markets are closed | 36.5 (32.3, 41.0) | 555 |
| **Proportion of household income lost during the pandemic (N = 1122)** |  |  |
| No income lost | 14.4 (12.3, 16.6) | 165 |
| 1-25% | 17.5 (14.6, 20.7) | 192 |
| 26-50% | 54.4 (51.3, 57.4) | 607 |
| 51-75% | 9.2 (1.7, 11.2) | 104 |
| 76-100% | 4.6 (3.5, 6.1) | 54 |
| **Percent reduction in household expenditures during the pandemic (N = 1122)** |  |  |
| No reduction | 36.3 (33.2, 39.6) | 415 |
| 1-25% | 23.2 (19.4, 27.4) | 257 |
| 26-50% | 35.7 (32.9, 38.6) | 400 |
| 51-75% | 3.9 (2.9, 5.3) | 41 |
| 76-100% | 0.89 (0.44, 1.8) | 9 |
| **Relative ability to access health care now compared to before the pandemic (N = 1121)** |  |  |
| Easier | 0.40 (0.15, 1.09) | 8 |
| No change | 47.0 (44.0, 50.0) | 544 |
| Somewhat harder | 37.4 (34.6, 40.2) | 413 |
| Much harder | 4.8 (3.7, 6.1) | 48 |
| Undecided | 10.0 (7.5, 13.1) | 108 |

The mean food consumption score was 60.9 (95% CI: 59.7, 62.3) (Table 2). Households consumed rice daily and meat and vegetables an average of 3.0 and 4.8 days per week, respectively. On average, children consumed 4.21 (95% CI: 3.95, 4.18) food groups in the day prior to the survey, corresponding to 62.5% (95% CI: 59.1, 65.8) of children that met the minimum DDS requirement. Women consumed an average of 5.38 (95% CI: 5.25, 5.51) food groups, corresponding to 67.7% (95% CI: 64.4, 70.9) meeting her minimum DDS. Compared to 2017, households in 2020 demonstrated significantly (p < 0.05) lower dietary diversity and household food security. In 2017, 76% of women and 69% of children met their minimum dietary diversity score, and the average CSI for households was 0.7 points lower. There was no change in household FCS from 2017 to 2020.

**Table 2.** Model coefficients representing difference in food security indicator between households who self-reported that it is harder to access food during the pandemic compared those who reported no change/easier; and those who decreased spending during the pandemic and those who did not. Adjusted models for households control for household ethnicity, household size, education level of mother and the head of household, and district. Adjusted models for mothers include additionally mother’s age. Adjusted models for children include additionally child’s age and sex. FCS = food consumption score; CSI = coping strategy index; DDS = dietary diversity score. Lower values for FCS and DDS and higher values of CSI indicate greater food insecurity.

|  | Model coefficients |  |  |  |  |
| --- | --- | --- | --- | --- | --- |
|  | Harder to access food during the pandemic |  | Decreased expenditures during the pandemic |  | Population mean (95% CI) |
|  | Crude difference (95% CI) | Adjusted difference (95% CI) | Crude difference (95% CI) | Adjusted difference (95% CI) |  |
| FCS | -3.36 (-5.42, -1.29)* | -2.74 (-4.92, -0.55)* | -6.53 (-8.23, -4.79)* | -5.24 (-7.05, -3.42)* | 60.9 (59.7, 62.3) |
| CSI | 0.07 (-0.86, 0.99) | 0.36 (-0.65, 1.37) | 0.83 (-0.07, 1.74) | 1.32 (0.40, 2.25)* | 3.6 (3.1, 4.1) |
| DDS (child) | -0.21 (0.41, -0.01)* | -0.20 (-0.41, 0.01) | -0.20 (-0.38, -0.02)* | -0.10 (-0.29, 0.09) | 4.14 (4.04, 4.24) |
| DDS (mother) | -0.15 (-0.40, 0.01) | -0.10 (-0.33, 0.12) | -0.08 (-0.28, 0.12) | 0.07 (-0.13, 0.26) | 5.38 (5.26, 5.51) |

\*represents statistical significance at $p < 0.05$

The distribution of both household food security indicators differed by whether or not households found it harder to access food during the pandemic (Figure 1). Among households who found it harder to meet their food needs during the pandemic, there was greater density of lower FCS (indicating worse food security) and higher CSI (indicating worse food security) compared to those who experienced no change. These relationships between household FCS and access to food during the pandemic were also seen in multivariate regression analyses (Table 2; Figure 2). Adjusting for ethnicity of the household, size of the household, district, and education level of the mother and head of household, we estimated that the average food consumption score among households who found it harder to meet their food needs was 2.74 points lower (95% CI: 0.55, 4.92) than the average food consumption score among households who experienced no change (Figure 2). This is roughly equivalent to consuming vegetables nearly three fewer times per week, or consuming rice one less time per week. The household coping strategies index among households who had a harder time meeting their food needs was higher, indicating lower food security, but not significantly so. Dietary diversity scores for women and children were lower among households who had more difficulty meeting their food needs during the pandemic, but not significantly so in adjusted analyses.

**Figure 1.**
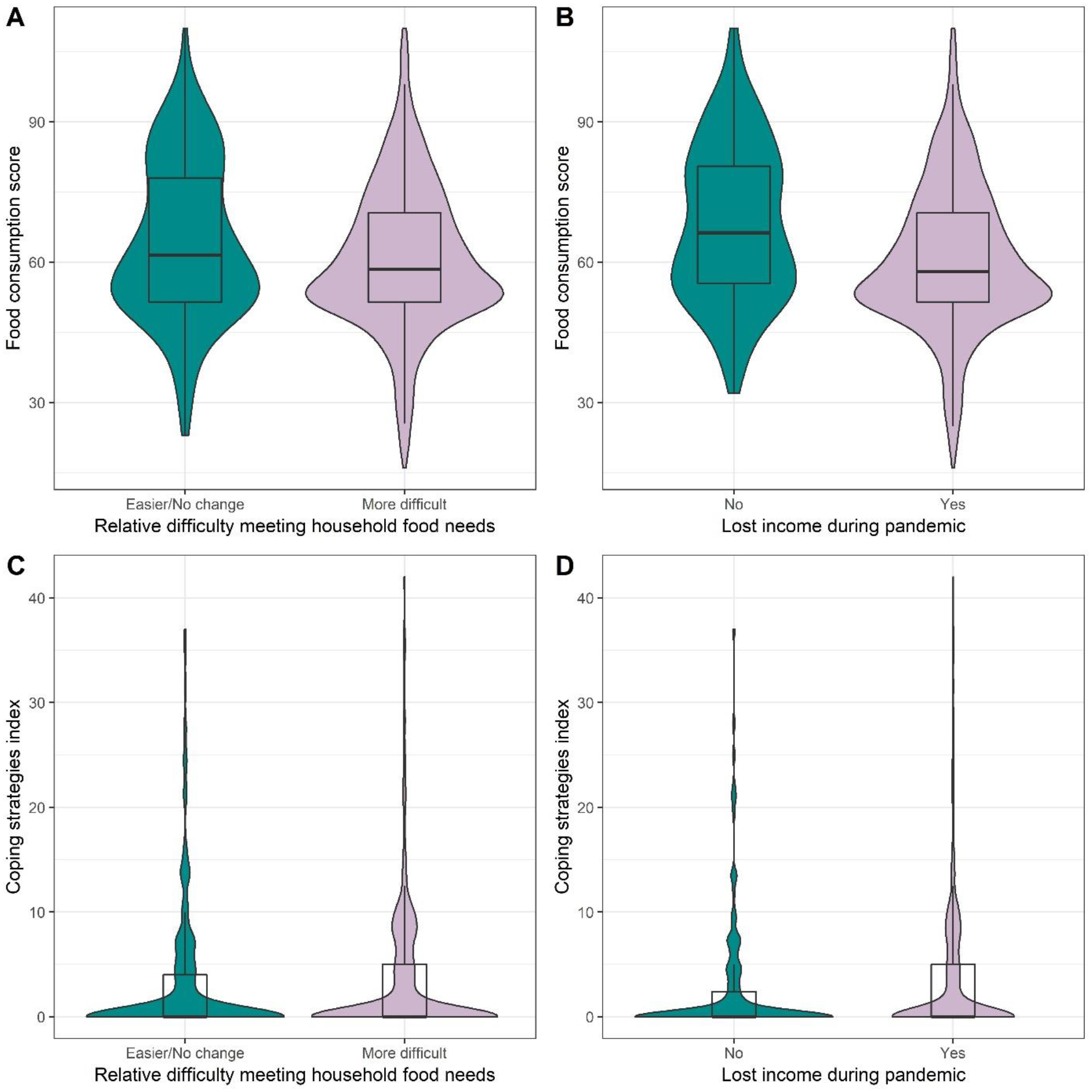
Violin plot showing distribution of two household food security measures, together with their median and interquartile range (IQR). Household food security was measured through food consumption score (FCS) (A, B) and coping strategies index (CSI) (C, D). Food insecurity is associated with low FCS and high CSI.

**Figure 2.**
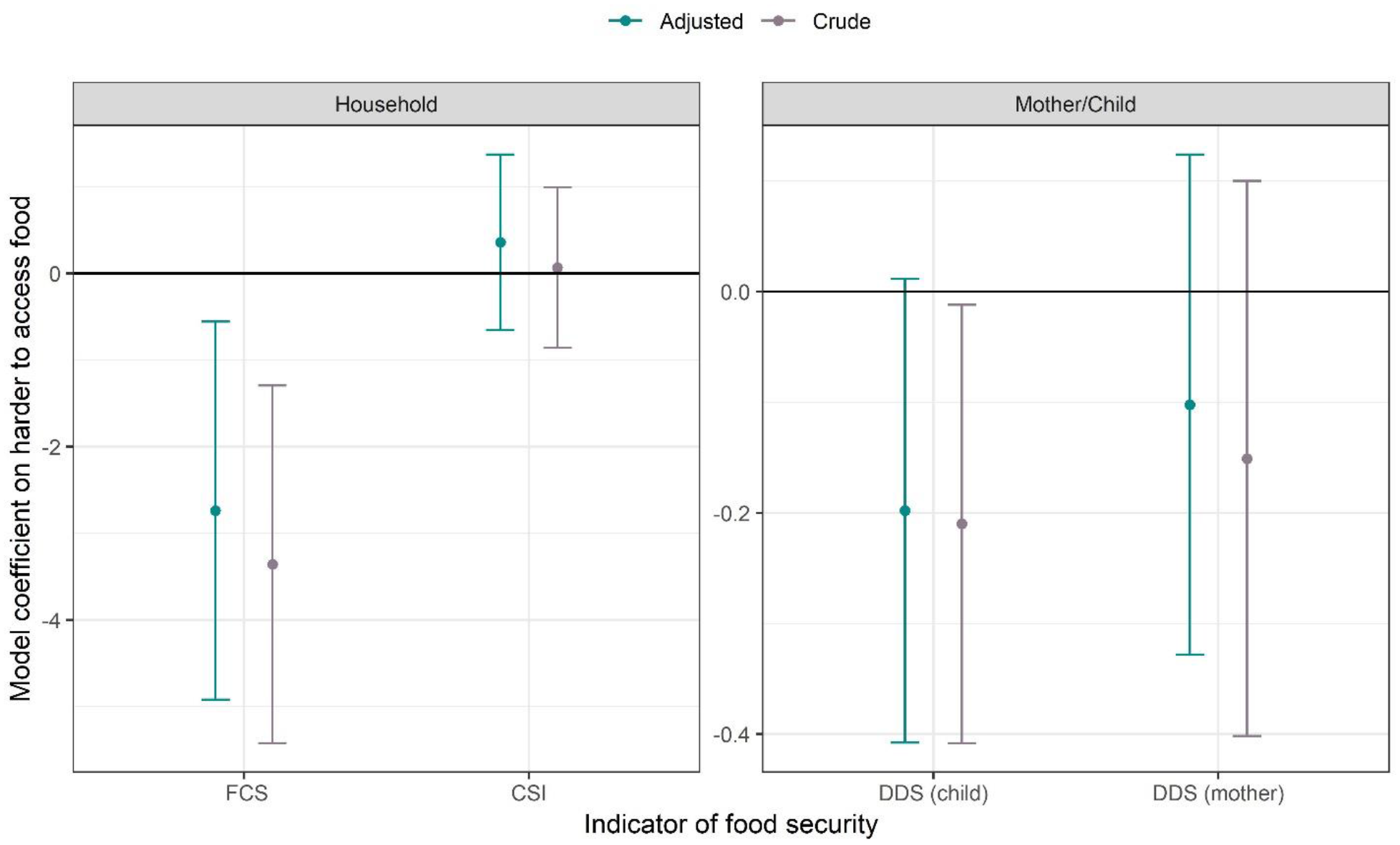
The difference in mean of food security indicator among households who had a harder time meeting their food needs during the pandemic compared to those who did not. Vertical bars represent 95% confidence intervals. Adjusted models for households control for household ethnicity, household size, education level of mother and the head of household, and district. Adjusted models for mothers include additionally mother’s age. Adjusted models for children include additionally child’s age and sex. FCS = food consumption score; CSI = coping strategy index; DDS = dietary diversity score. Lower values for FCS and DDS and higher values of CSI indicate greater food insecurity.

### Resiliency to food insecurity

We estimated the percentage of a household’s food sources in the past week that was self-produced (e.g., farmed, fished, hunted, gathered). On average, families met 42% of their food needs through self-production (interquartile range: 27%, 57%). Commonly self-produced foods included: insects, aquatic animals other than fish, mushrooms, and roots (Figure 3). Over half of households also self-produced rice and vegetables, and about one quarter self-produced fish, meat, and fruits. We found that households who derived a greater proportion of their food needs through homegrown methods were more resilient than families who purchased their foods. Adjusting for ethnicity of the household, size of the household, district, and education level of the mother and head of household, we estimated that the average percentage of food obtained from homegrown methods was 4.22% (95% CI: 1.28, 7.15%) lower among households who found it harder to meet their food needs compared to household who experienced no change. Persons who found it harder to meet their food needs during the pandemic also spent fewer hours per week fishing, gathering, or hunting, though the results were not significant.

**Figure 3.**
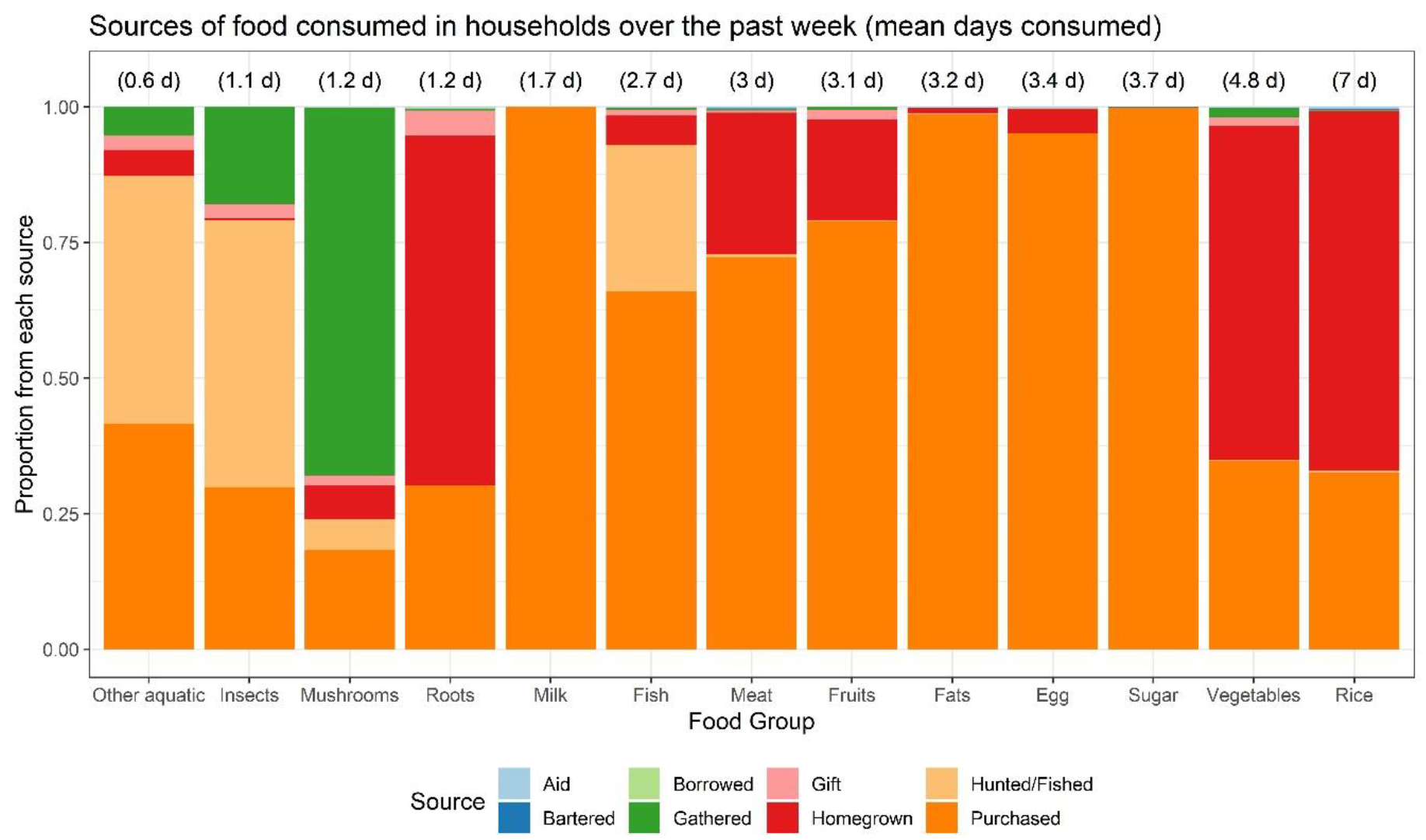
Proportional source of each food group consumed during the past week by households. Numbers in parenthesis above the bars indicates the mean number of days per week household consumed these food groups.

### Income and expenditures

Over 85% of the study population reported losing income during the pandemic, with the majority of respondents (54.4%, 95% CI: 51.3, 57.4%) reporting losing between 25-50% of their income. Households who reported declines in income were more likely to reduce spending, with the greater the reduction in income corresponding to greater reductions in household expenditures (Figure 4a). A weighted 23.3% reported reducing household expenditures by 1-25%, while 35.7% reported reducing expenditures by 25-50%. The distribution of both household food security indicators also differed by whether or not households lost income during the pandemic (Figure 1).

**Figure 4.**
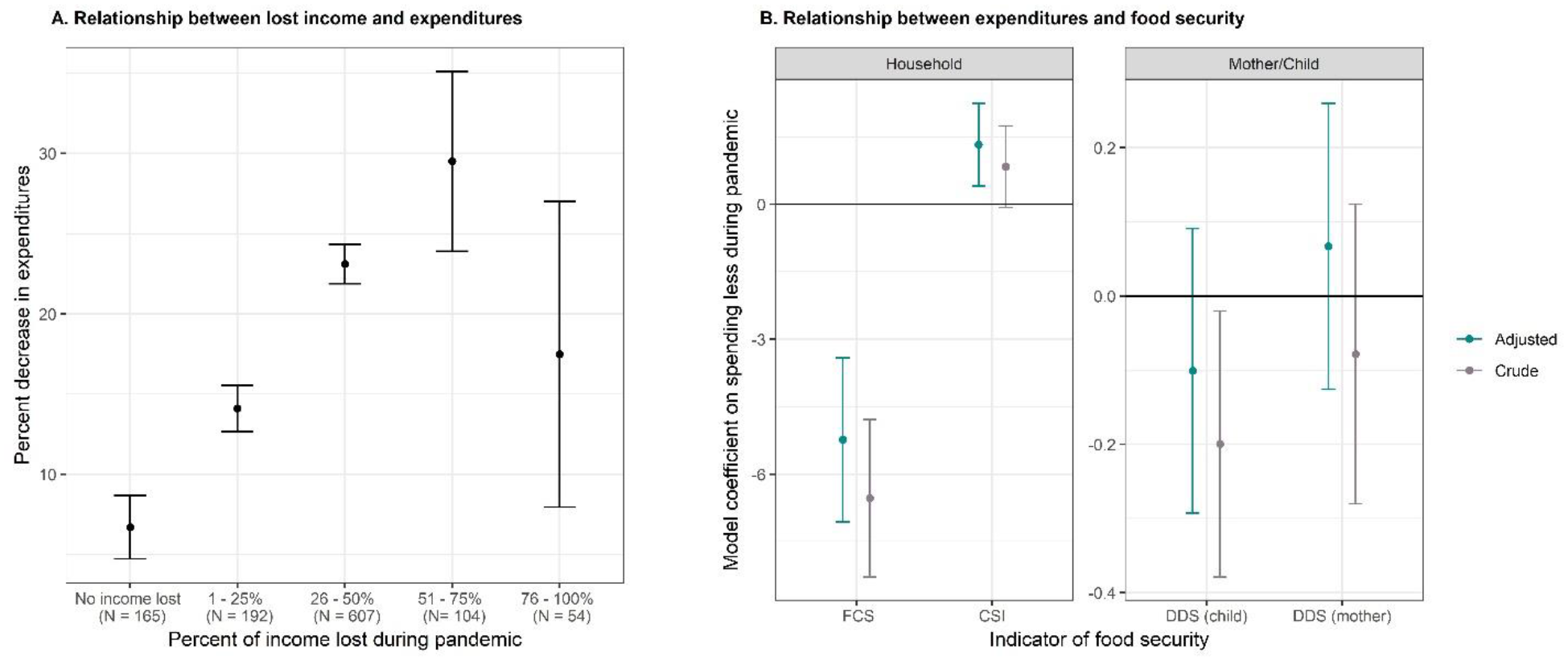
A) Mean decrease in expenditures reported, stratified by the percent reduction in household income. Vertical bars represent 95% confidence intervals. B) The difference in mean of food security indicator among households who reduced spending during the pandemic compared to those who did not. Vertical bars represent 95% confidence intervals. Adjusted models for households control for household ethnicity, household size, education level of mother and the head of household, and district. Adjusted models for mothers include additionally mother’s age. Adjusted models for children include additionally child’s age and sex. FCS = food consumption score; CSI = coping strategy index; DDS = dietary diversity score. Lower values for FCS and DDS and higher values of CSI indicate greater food insecurity.

Households who reduced expenditures during the pandemic had significantly decreased food security in adjusted analyses, as measured by the FCS, and significantly decreased food security in univariate analyses as measured by the FCS, CSI, and child’s DDS (Figure 4b, Table 2). In adjusted analyses, families who reported spending less during the pandemic had a household FCS that was 5.23 (95% CI: 3.41, 7.05) units lower, and a CSI that was 0.83 (95% CI: −0.07, 1.74) units higher than families who did not reduce spending. Dietary diversity scores for children were lower among households who had more difficulty meeting their food needs during the pandemic, but not significantly so in adjusted analyses.

### Access to health care

A weighted 37.4% (95% CI: 34.6, 40.2%) of individuals reported that it was somewhat harder to access healthcare compared to before the pandemic, while 4.8% (95% CI: 3.7, 6.1%) reported that it was much harder (Table 1). We identified 123 (11%) women and 557 (50%) children who had experienced fever, diarrhea, or respiratory infection in the two weeks prior to the survey. Of these, a weighted 69.7% (95% CI: 66.3, 73.0%) of children and 81.2% (95% CI: 73.3, 87.2) of women sought care from a health facility. We found no association between healthcare seeking behavior and relative ability to access health care.

### Sensitivity of undernutrition prevalence to small shocks in bodyweight

We did not find any difference in WAZ or WHZ scores among children from households who experienced greater difficulty meeting their food needs or among children from households who lost income or reduced spending. We examined the change in the proportion of children classified as wasted or underweight under simulated shocks in which bodyweight decreased by 0.5% and 1%. In the study population, we observed a prevalence of wasting of 4.5%. If bodyweight were to decrease by 0.5% or 1%, we estimated a prevalence of wasting of 6.2% and 6.6%, respectively, in our population (Figure 5). In other words, a decrease in bodyweight by 0.5 – 1% would be associated with a disproportionate increase in wasting of 1.7 – 2.1 percentage points in our study population. Similarly, we observed a prevalence of underweight of 18.2%. If bodyweight were to decrease by 0.5% or 1%, we estimated a prevalence of underweight of 19.0% and 20.5%, respectively. Therefore, a decrease in bodyweight by only 0.5 – 1% would be associated with an increase in underweight of 0.8 – 2.3 percentage points in our study population.

**Figure 5.**
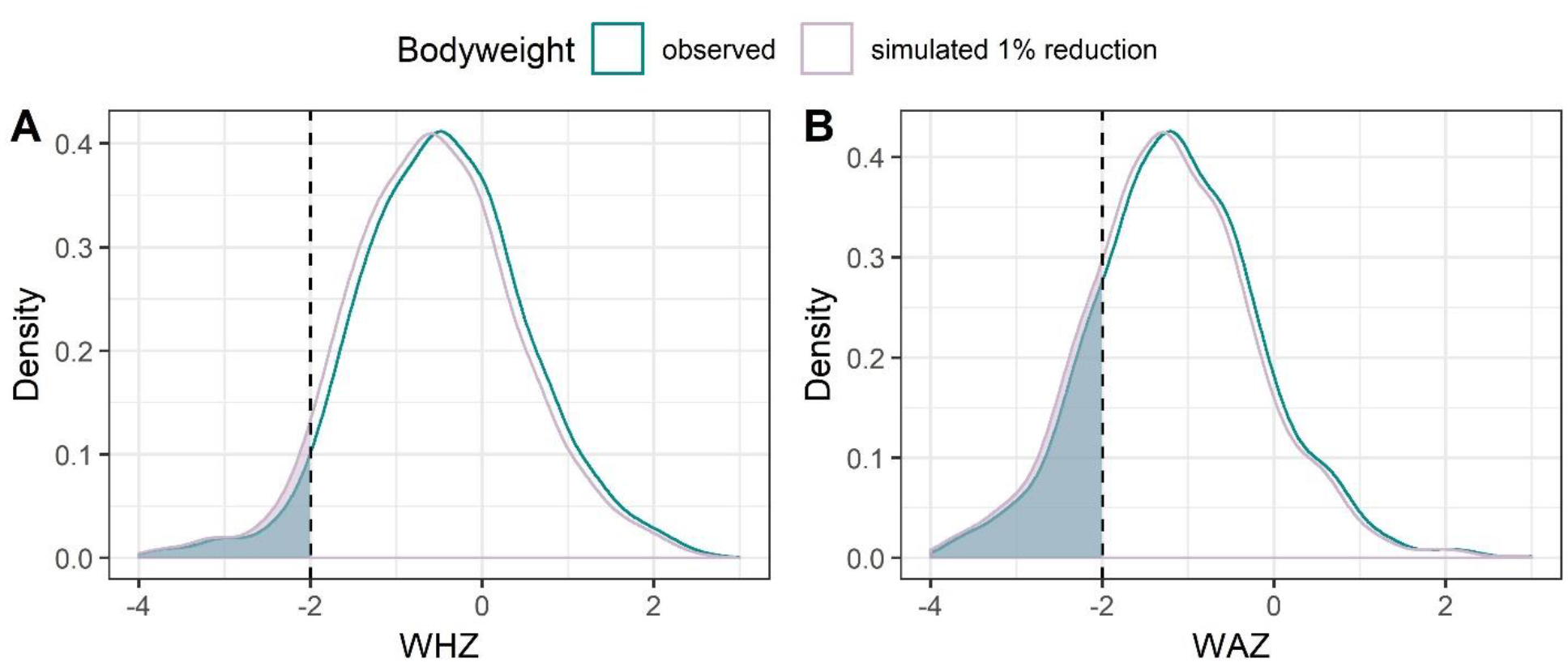
WHZ and WAZ curves among children under 5 under observed (cyan) conditions and under a simulated shock in which body weight reduces by 1% (pink). Area shaded to the left of - 2 represents the proportion of children classified as wasted or underweight, respectively.

## Discussion

In a rural setting in Lao PDR with low documented COVID-19 transmission and high dependence on tourism, we found prevalent loss of income and increased difficulty in meeting household food needs following the start of the COVID-19 pandemic and a national border closure. In our household survey, we found that nearly four-fifths of the study population reported that it was harder to meet their family’s food needs during the pandemic, with the most common reason being increases in food prices; indeed, families reported that the proportion of their household expenditure on food had doubled since baseline in 2017. At the same time, we found that over 85% of the study population reported losing income during the pandemic, with over half of respondents reported losing between 25-50% of their income. Respondents who reported losses in income and and/or reported greater challenges meeting their food needs had small, but significant declines in household food security, as measured by the food consumption score and coping strategies index. Nevertheless, the small differences in food security indicators suggests that people in this population may have been able largely able to protect their consumption without heavy reliance on negative coping strategies, despite some deterioration. Self-production of food via farming, hunting, fishing, or fathering is common in this population, accounting for 42% of food consumed. Our study found that individuals who derived a greater proportion of the food from self-produced means were more resilient to pandemic-associated shocks.

Our results support a limited, but growing, body of empirical data that suggests wide scale difficulty in meeting food needs and pervasive loss in income associated with the pandemic. In Kenya, surveys administered before and after the COVID-19 lockdown found that 52% of the population changed their dietary habits, most commonly via reductions in meat, dairy, and bread [30]. Nearly all (95%) of respondents reported loss of income during the pandemic, with 88% finding that the resulting income was insufficient to meet food needs. Over one third also attributed changes in food consumption to lower food availability [30]. An interrupted time series analysis in Bangladesh found that median incomes fell from US$212 to $59 during a two-month stay at home order, while the proportion of families living on less than $1.90 per day rose from 0.2% to 47.3% [31]. In this study, the proportion of households classified as moderately or severely food insecure rose from 5.6% and 2.7%, respectively, to 36.5% and 15.3% [31]. Finally, in a Save the Children global survey, 85% of families living in Asia reported income loss, with a strong negative association between income loss and dietary diversity [21]. No study has yet to be published from Lao PDR, but an unpublished household survey in Phongsaly Province, another rural province, found that 46% of households reduced their expenditures, and 24% took out loans to buy food (personal communication).

Randomized control trials demonstrate that improved access to proper nutrition can improve WAZ and WHZ Z-scores [32–34]. We examined theoretical implications of a decrease in bodyweight on undernutrition prevalence, finding that a decrease in bodyweight of only 0.5 – 1% would be associated with a larger percentage point increase in wasting (1.7 – 2.1 percentage points) and underweight (0.8 – 2.3 percentage points) in our study population. While LMICs have seen progress in reducing prevalence of wasting and underweight, yearly reductions are small. Analysis of DHS data collected between 1990 and 2012 from 36 LMICs found that, on average, the prevalence of wasting decreased by 0.07 percentage points per year [35], while in Lao PDR, the prevalence of underweight decreased by an average of 1.1 percentage points per year between 2012 and 2017 [14, 36]. This suggests that even small effects of COVID-19 on food security, and thus bodyweight, could undo years of progress. This echoes findings from a study conducted in India and is likely generalizable to many LMICs where there is a high prevalence of undernutrition [27]. At the same time, we did not observe a difference in the WAZ or WHZ scores between children whose household reported greater difficulty meeting food needs and those who did not, nor did we see a difference in child dietary diversity score between these groups in multivariate analyses. This may suggest that households in our study population prioritized maternal and child consumption patterns even as families struggled to meet food needs. All villages in the study population have been receiving interventions focused on sustainable behavioral change for maternal and child nutrition, so individuals in the population may have been more likely to prioritize the nutrition of these vulnerable populations.

Our study suggests possible interventions that might mitigate the effect of the pandemic on food security. We found that households who were more likely to experience no change in meeting food needs during the pandemic derived a greater proportion of their food needs through homegrown methods (as opposed to purchasing foods) as compared to households who found it more difficult to meet their food needs. Reducing reliance on food supply from other places or countries is recognized by others to be a means of reducing the impact of the COVID-19 pandemic on food insecurity. Farm-system-for-nutrition approaches have been suggested as one solution, in which location-specific farm systems that integrate arable farming, horticulture, backyard farming, and animal farming [37]. The FAO advocate for improving the resilience of local food systems by facilitating access to locally produced food, shortening the supply chain by promoting direct purchase from local producers, and promoting urban or backyard gardens that also offer financial and environmental co-benefits [38].

Our study also identified that loss of income and higher food prices are among the most important reason households are less able to meet their food needs. As such, social safety net programs may be particularly suited to addressing the challenge of food insecurity [7, 39, 40]. A randomized control trial in Colombia in March 2020, at the start of a national quarantine, found that 90% of families randomized to an arm that received cash transfers of $19 every 5-9 weeks spent the cash on food, which helped to offset the effects of the pandemic on food insecurity in the treatment arm [41]. Other randomized control trials demonstrate reductions of severe food insecurity among those who received a cash transfer or a direct food transfer by nearly 25% [42, 43]. Systematic review and meta-analysis of 74 studies found that children from households who received cash transfers had reduced stunting by 2.5% and improved consumption of animal foods by 4.5% [44].

This study has limitations. First, the results of this survey may not be generalizable to other countries, particularly those with higher COVID-19 incidence and greater restrictions on within-country movement. At the time of the survey (November 2020), fewer than 50 cases had been reported in Lao PDR, and health systems were not experiencing the same overwhelming of capacity as in many other countries [45]. Additionally, while initial control measures limited local movement, these restrictions were largely relaxed by May 2020, seven months prior to the survey, with the main intervention remaining being strict border closure. We expect, therefore, that compared to other LMICs, the effects of food security and access to health care found in this study may be smaller than would be seen in other countries. At the same time, however, the effects of the pandemic on food security and income and expenditures may be seen more strongly in Luang Prabang as compared to other provinces within Lao PDR. As the province is home to the UNESCO World Heritage City of Luang Prabang, Luang Prabang province receives a greater proportion of its income from tourism as compared to other provinces [20]. Indeed, our survey found a greater proportion of household reduced expenditures (64%) compared to another, unpublished, survey in a different rural province, where 46% of households reduced expenditures (personal communication). As mentioned, households in the study population had been receiving educational messaging regarding the importance of maternal and child malnutrition, so may have prioritized meeting the needs of mothers and children even as their struggled to meet the families’ food needs. Thus it is possible that other areas may have seen more dramatic declines in maternal and child nutrition. Moreover, the results of the survey may not be generalizable to larger, more urban areas. Finally, the relationships with FCS may not be generalizable to other areas with different dietary patterns. The mean FCS in our study was 60.9, well above the generic cut off of ≥35 for an acceptable score. While diversity of foods consumed was low, consumption of staples and meat/fish/insects was high, and these food groups are given large weights in calculating the weighted mean.

Another limitation of our study relates to recall bias. Because control measures were first implemented in March 2020, and we implemented this survey in November 2020, there could be substantial recall bias, as participants are asked to compare ability to meet food needs, ability to access health care, and income and expenditures to a time period that extended 8 months prior up until the current time. The ideal observational research design would be to compare our estimates of food security and malnutrition to repeated estimates taken longitudinally, leading up to just prior to the pandemic. While we lack data from just before the pandemic, we have data from household surveys in the region collected in 2017. Estimates of food insecurity and the prevalence of children underweight and wasted from 2020 are higher than estimates from 2017, while estimates of dietary diversity from 2020 are lower than estimates from 2017. However, because changes in indicators between 2017 and 2020 cannot be attributed to the effects of the pandemic alone, we do not emphasize 2017 data here.

## Conclusion

Lao PDR’s early efforts to control the spread of COVID-19 have been successful, with fewer documented cases to date relative to neighboring countries. Nevertheless, the effect of the pandemic on food security in livelihoods in LMICs may be severe, and the second wave of cases, and associated lockdown measures, in April 2021 demonstrates that the threat of continued food security remains present. Increasing self-sufficiency through local food production, and/or supporting incomes via social safety nets such as cash transfer programs, may mitigate some of these effects. As control measures to curb the transmission of COVID-19 continue, and as outbreaks occur intermittently with concomitant restrictions on movement, further study may be useful to understand what coping strategies people are using so that government and agencies can support the resilience of households in the long term.

## Data Availability

Data is owned by the Luang Prabang Provincial Health Department and permission has been granted for its use.

## List of abbreviations

LMICs: low- and middle-income countries
FAO: Food and agriculture organization
FCS: food consumption score
CSI: coping strategies index
DDS: dietary diversity score
HAZ: height-for-age Z-score
WAZ: weight-for-age Z-score
WHZ: weight-for-height Z-score

## Declarations

### Ethics approval and consent to participate

Data were collected by the Lao Provincial Health Department as part of routine, non-research public health activities. We requested and obtained data from the Lao Provincial Health Department. Ethical clearance for secondary data analysis was obtained from the Research Ethics Committee in the University of Health Sciences within the Lao Ministry of Health and Committee for the Protection of Human Subjects within University of California, Berkeley (protocol ID: 2021-05-14365).

### Competing interests

HC, AV and KK, were or are currently employees of Save the Children, International. Save the Children supports a government led Primary Health Care Program in Luang Prabang which includes nutritional interventions.

### Funding

The survey was funded from the grants received by Save the Children Japan from Takeda Pharmaceutical Company Limited Global CSR Partnership.

### Authors’ contributions

PC, HC, and JRH conceptualized the research. PC and HC assisted in data collection. PC and JRH analyzed the data. HC and JRH wrote the manuscript. AV and KK lead the Save the Children health program in Luang Prabang and the Vientiane country office, respectively. All authors edited and read the manuscript.

## Acknowledgements

We are grateful to Lilly Schofield and Yasir Arafat for their inputs on COVID related questions in preparing the survey and their review of the manuscript. We are grateful for the team of data collectors and supervisors who collected the data, to our study participants for their time and investment in the survey, and to the Luang Prabang Provincial Health Department for their continued partnership.

## Supplemental Info: Survey tool

### Endline Interview Questionnaire – 2020 Health and Nutrition Assessment

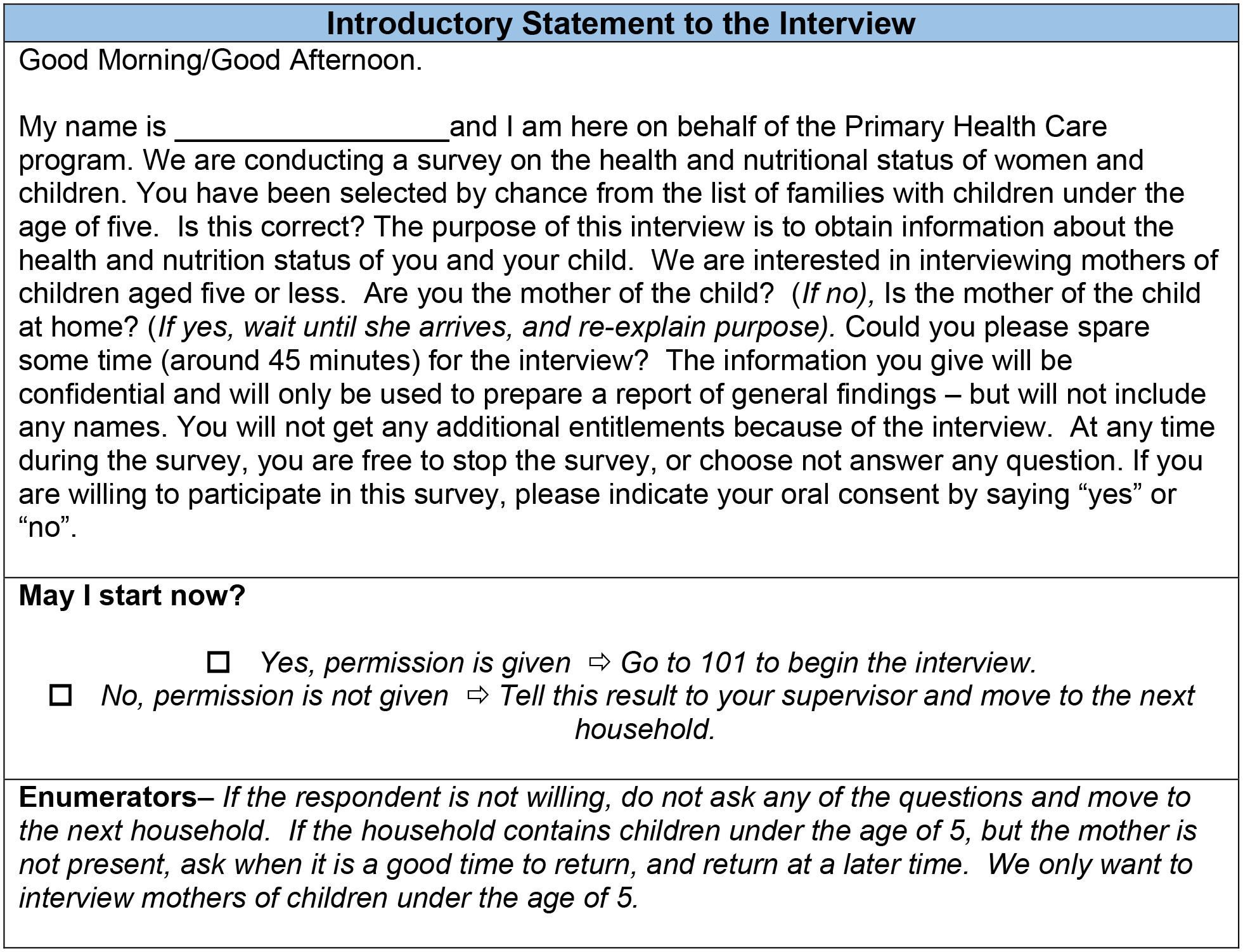
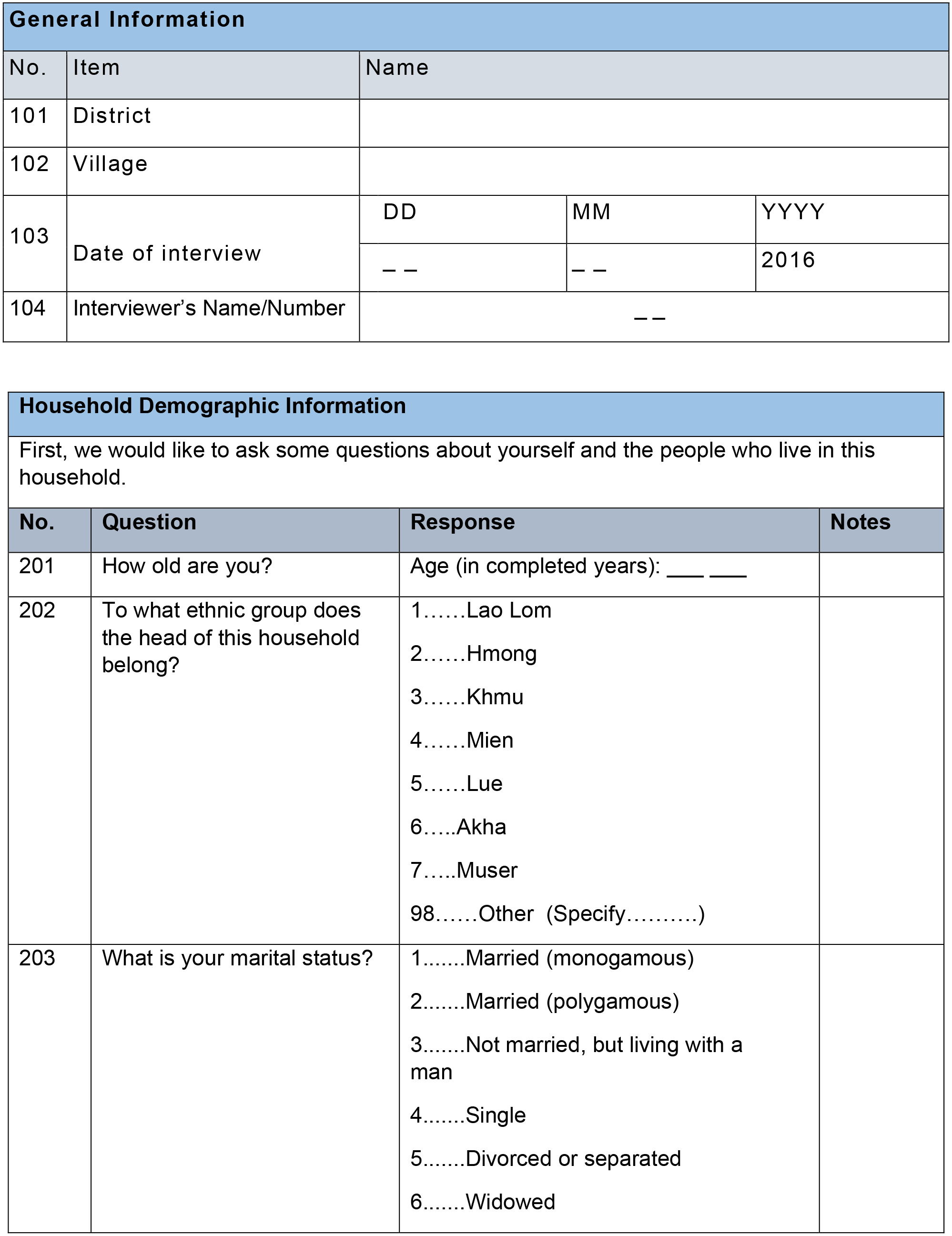
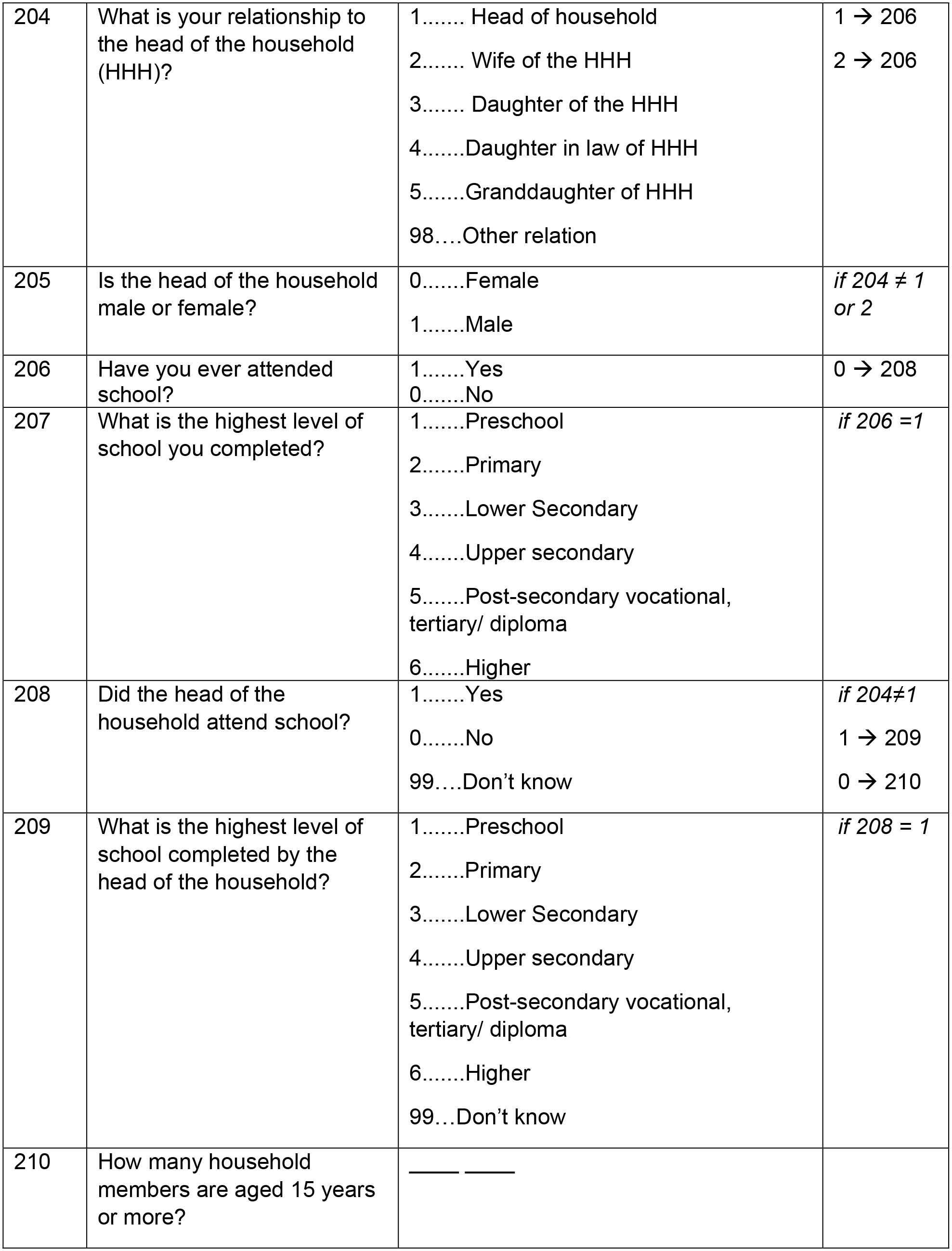
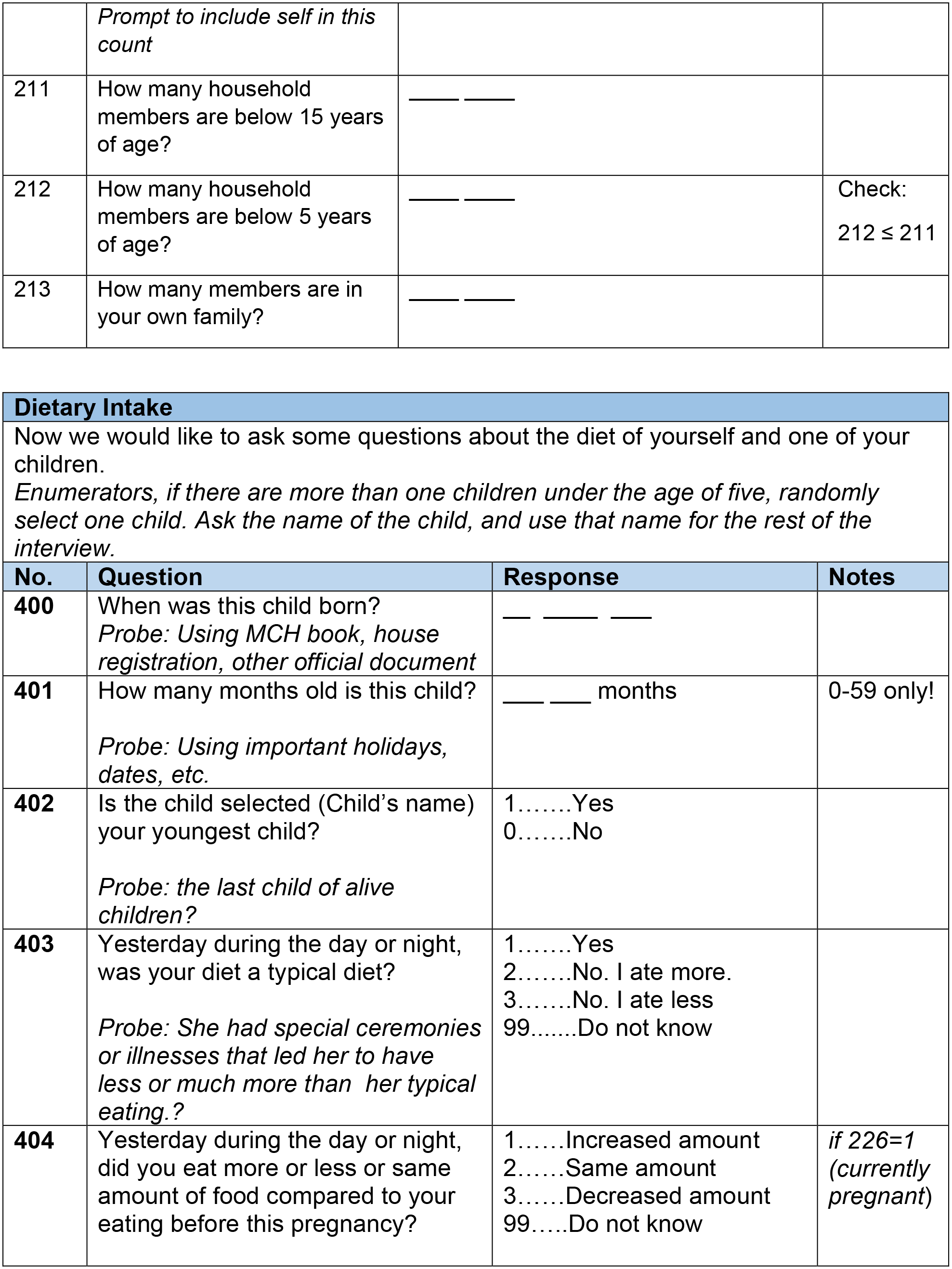
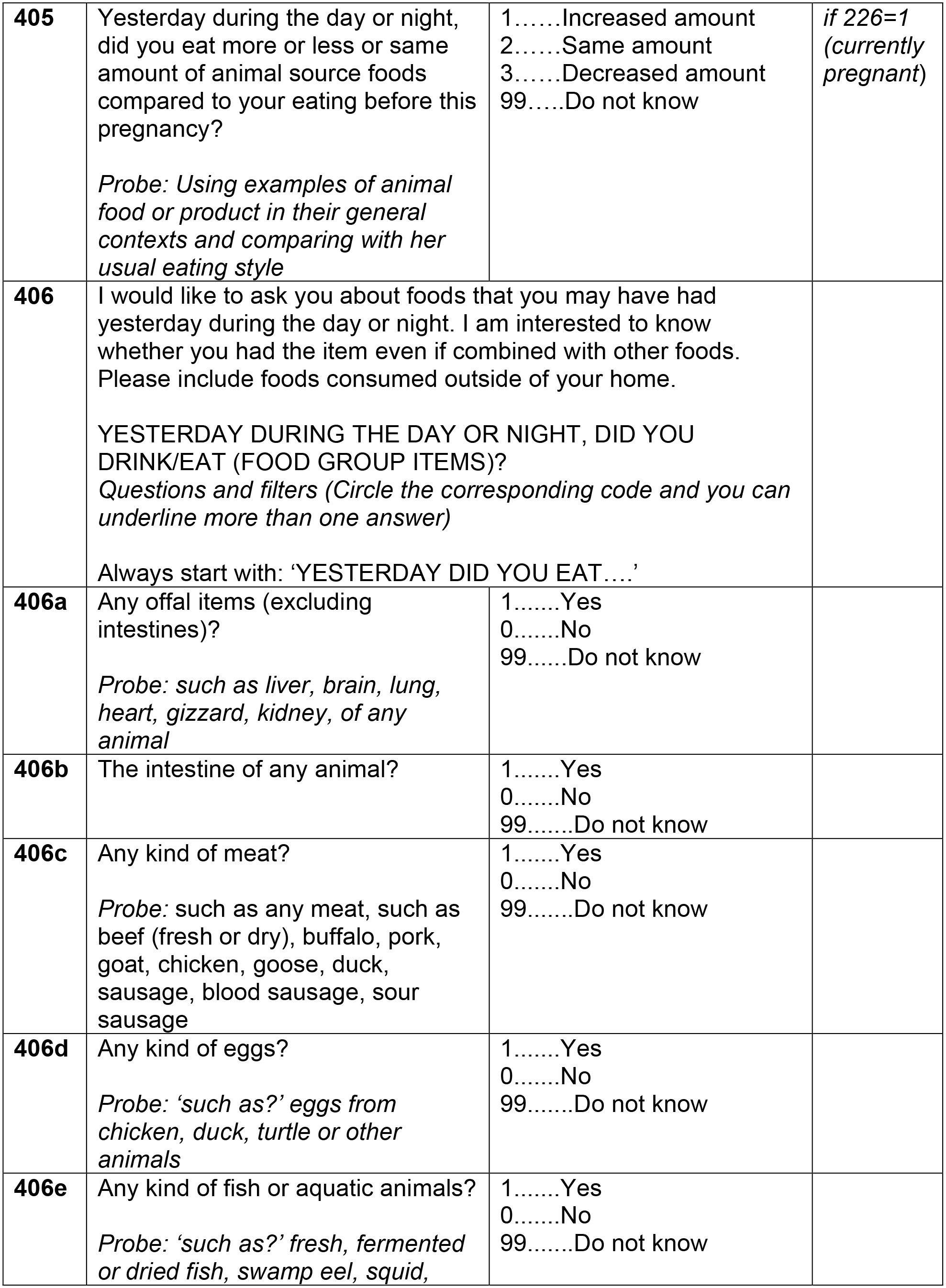
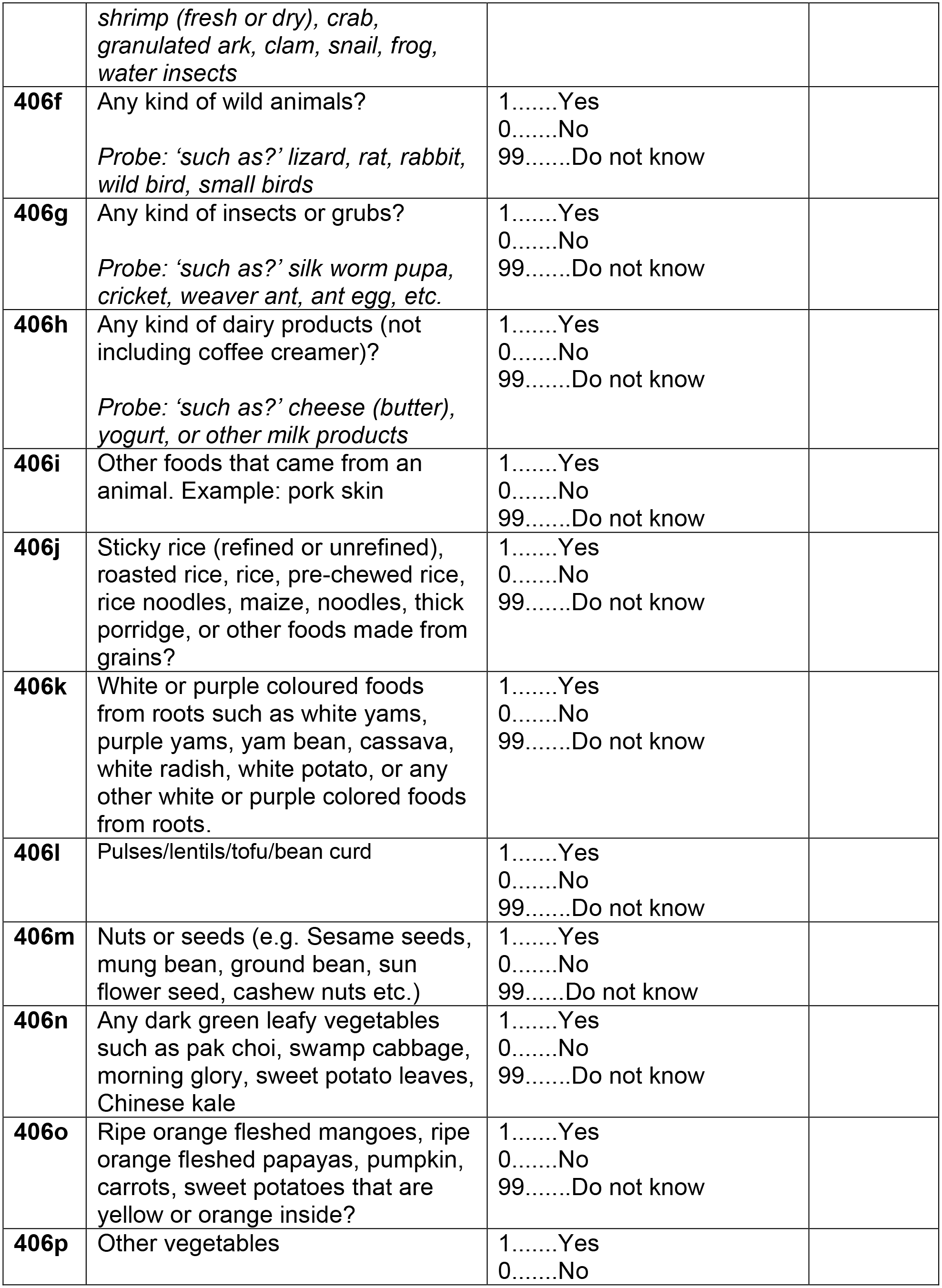
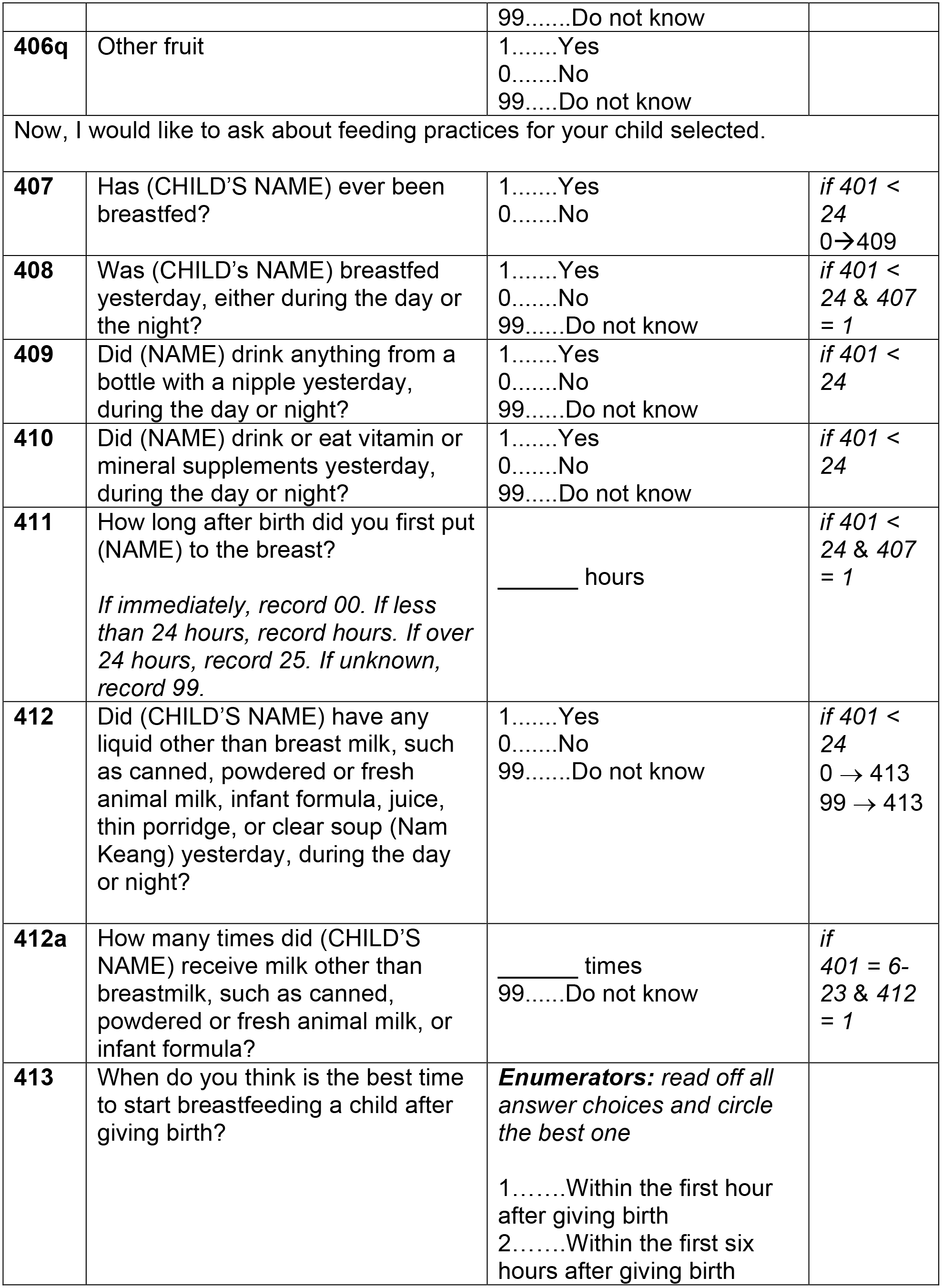
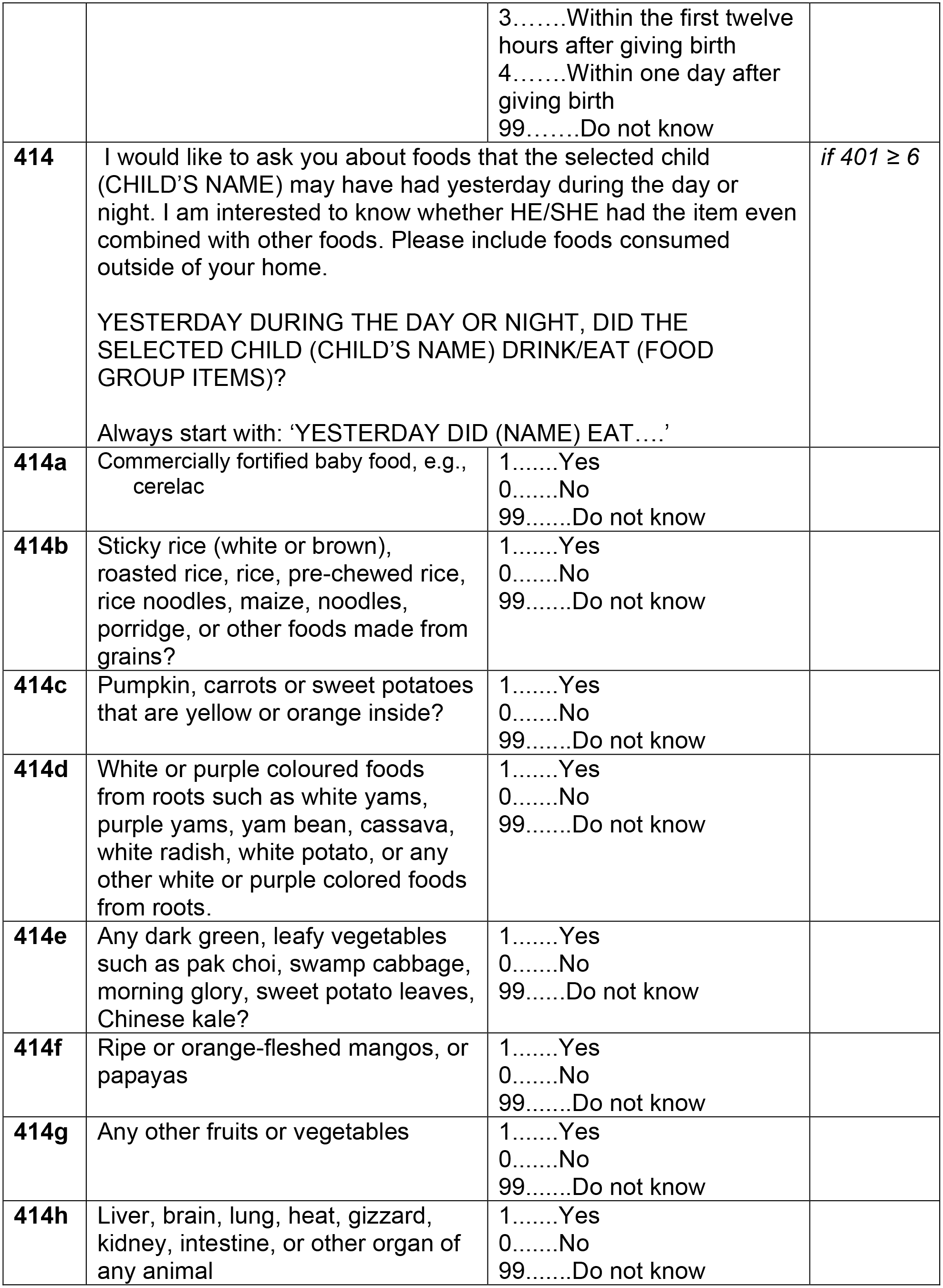
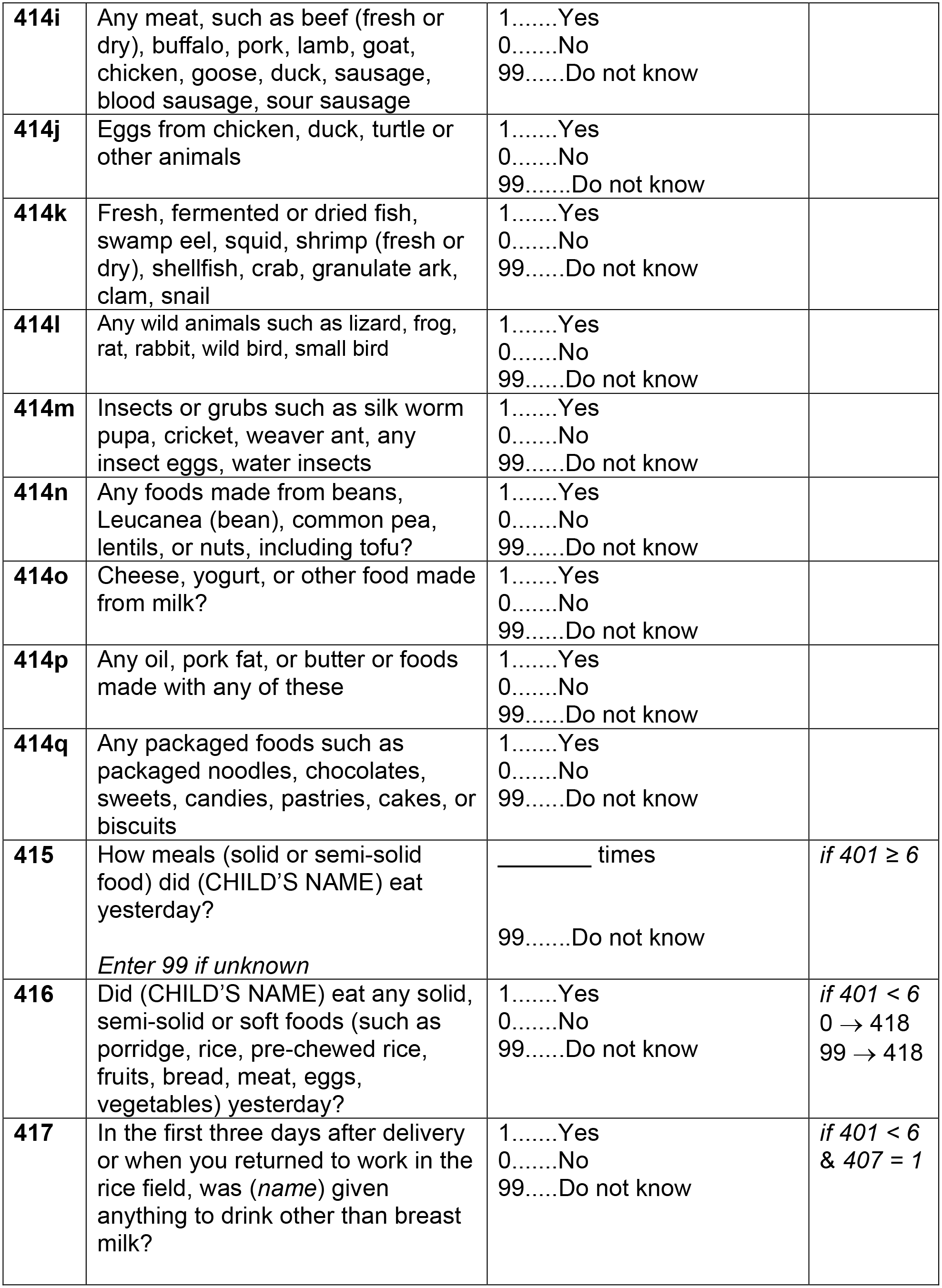
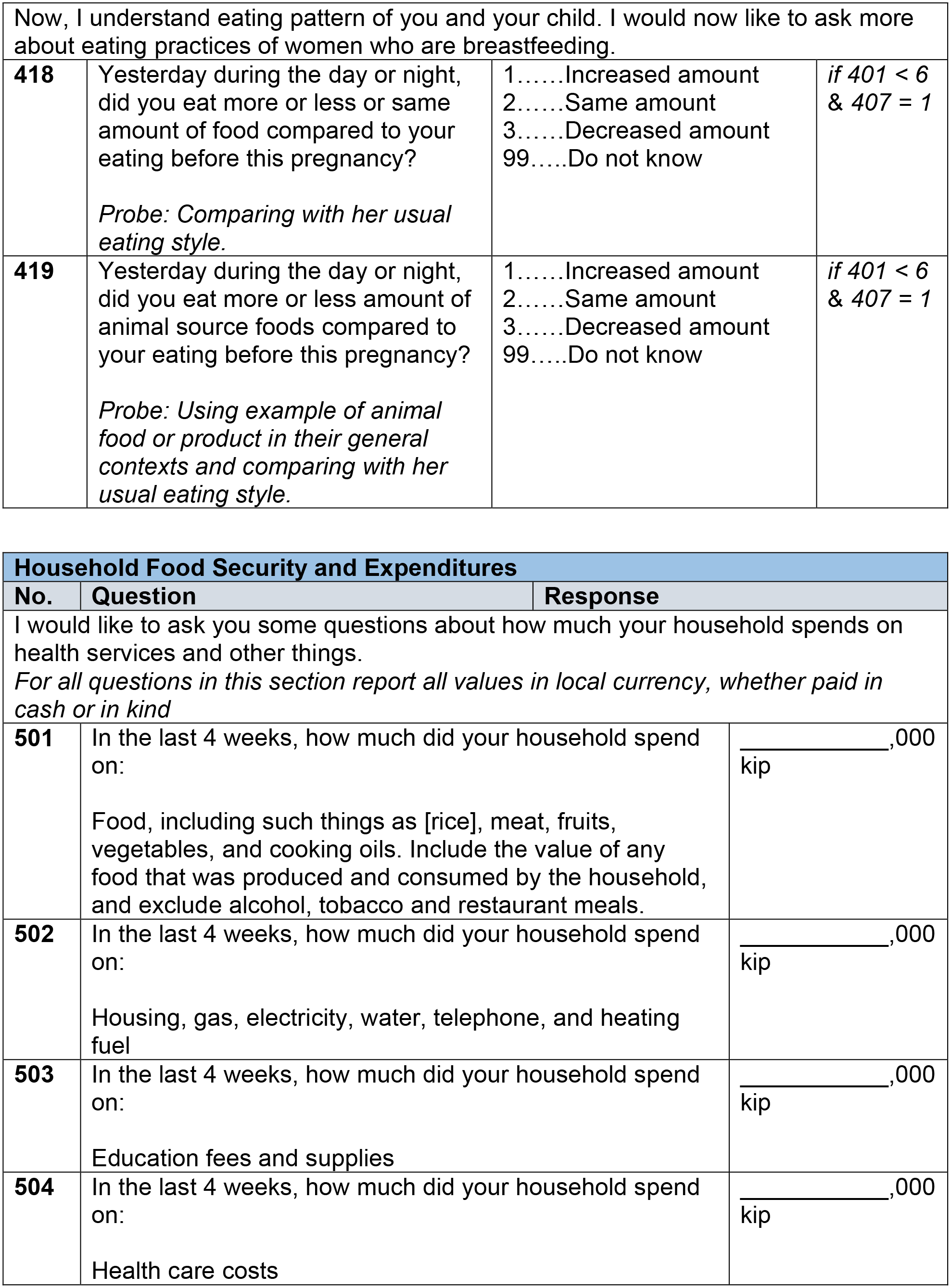
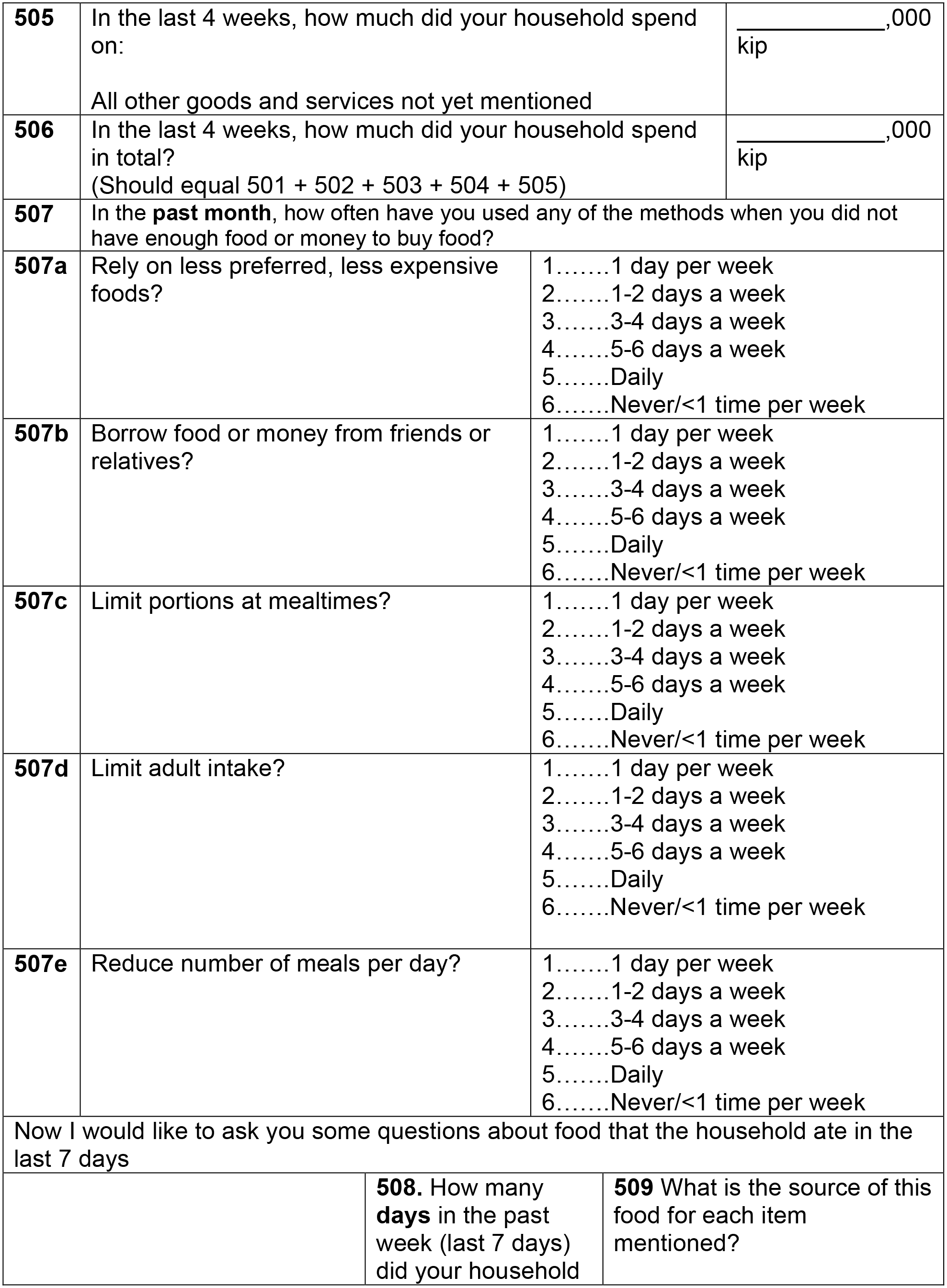
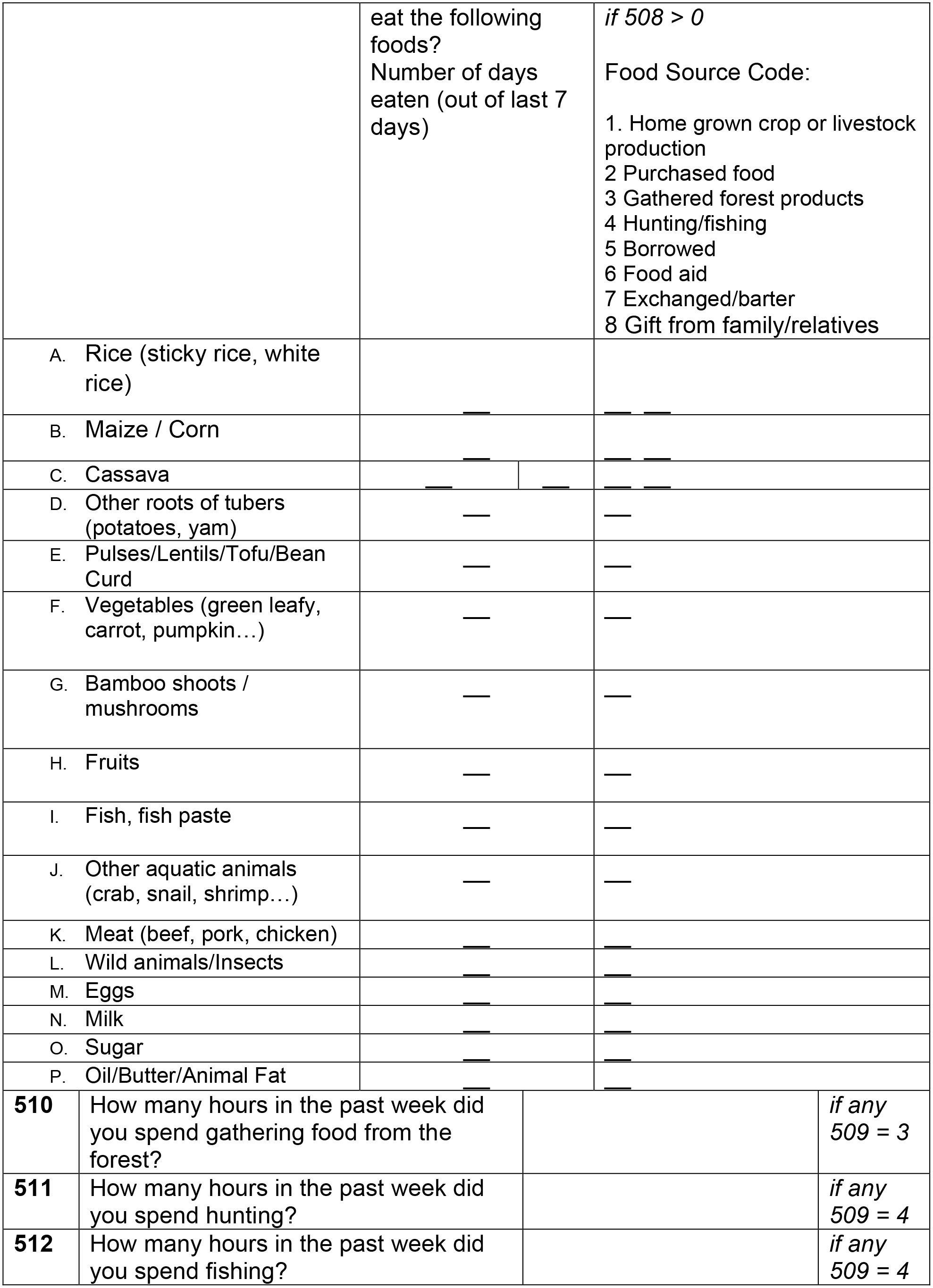
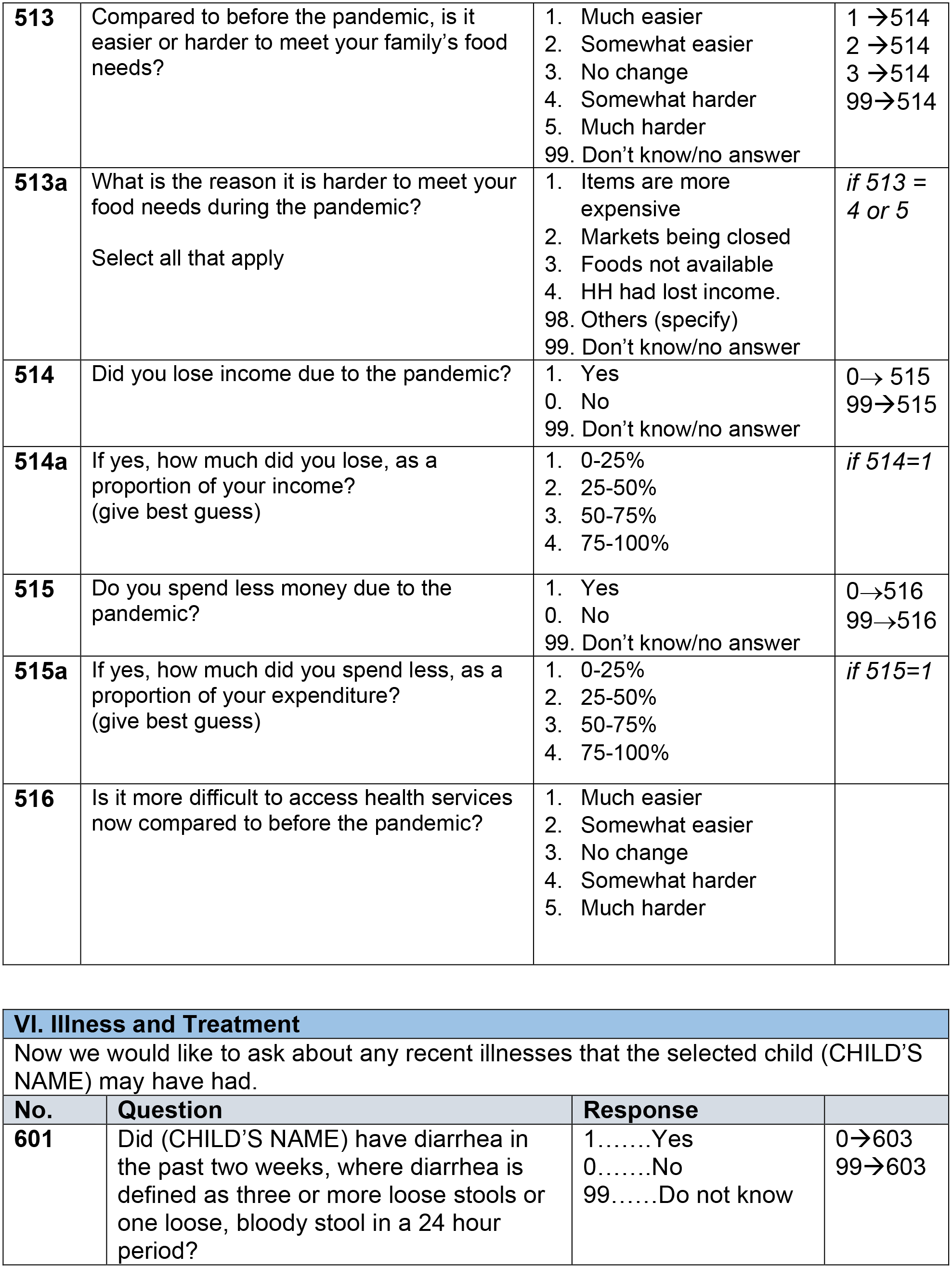
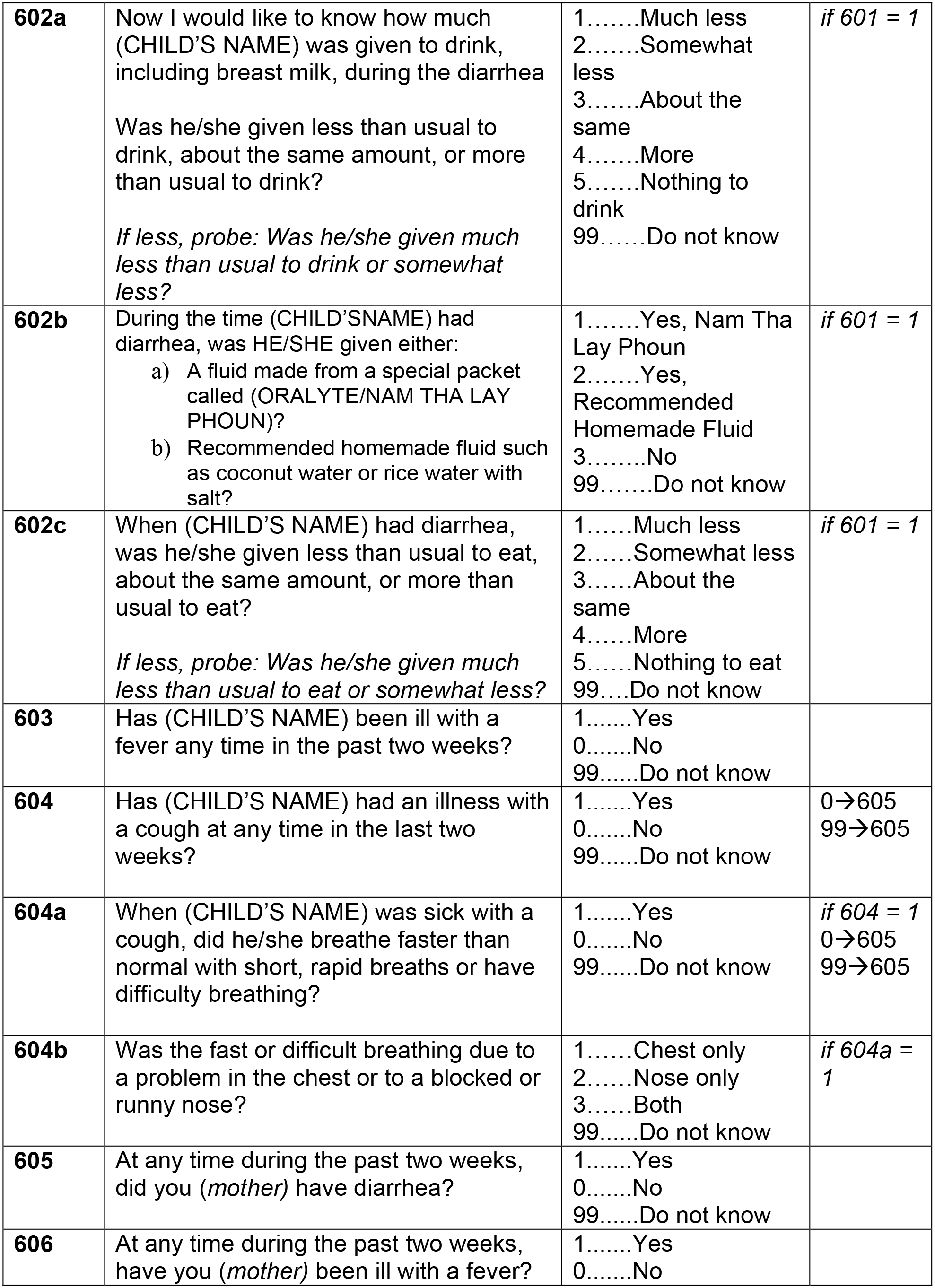
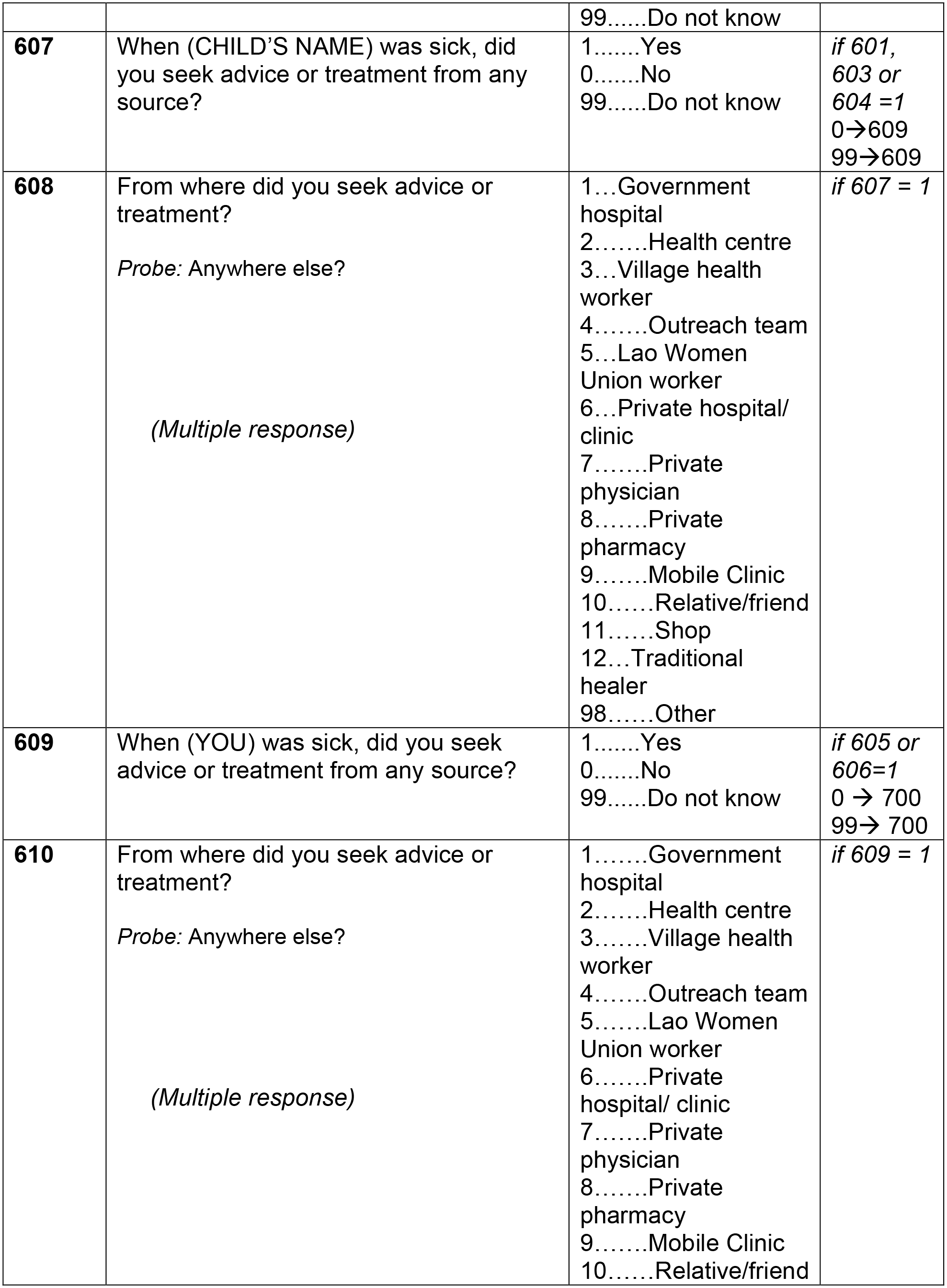
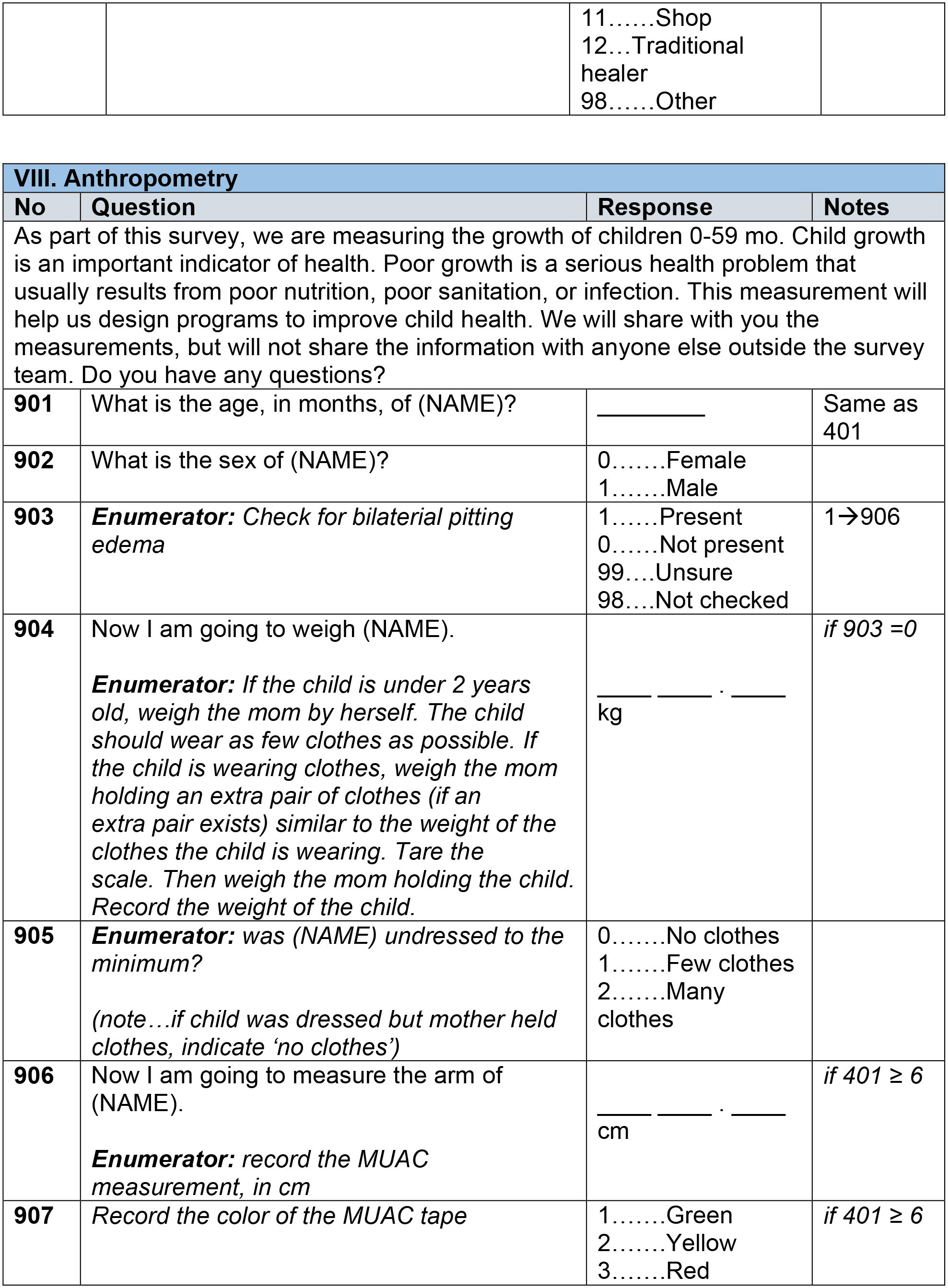
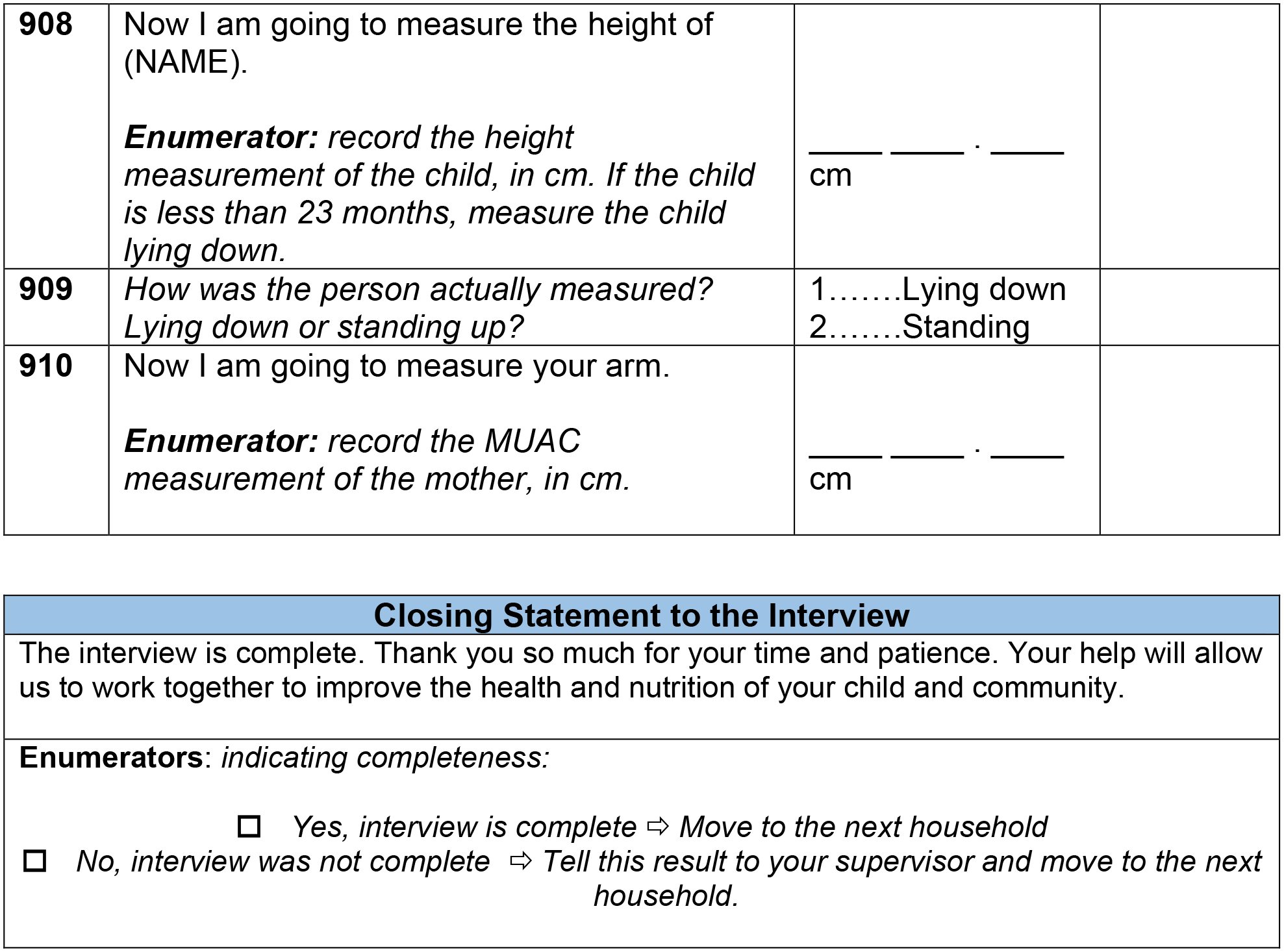

